# Exploring multifaceted mental resilience from the perspectives of sleep and circadian rhythm in healthy adults

**DOI:** 10.64898/2026.09.25.26363918

**Authors:** Chris Xie Chen, Ran Wang, Forrest Tin Wai Cheung, Ngan Yin Chan, Tim Man Ho Li, Joey Wing Yan Chan, Jihui Zhang, Wai Kai Hou, Suk Yu Yau, Yan Liu, Tatia Mei Chun Lee, Shirley Xin Li, Yun Kwok Wing

## Abstract

**Background:** Emerging evidence suggested the role of sleep and circadian rhythm in the development of mental resilience, but the association of an integrated analysis of objective sleep and circadian measures with multifaceted mental resilience in healthy adults has been underexplored.

**Objective:** This study aims to explore the associations between multifaceted mental resilience and sleep and circadian rhythm in healthy adults.

**Methods:** A total of 156 healthy adults (Age: Mean (SD): 29.54 (5.28) years; 60.9% female) underwent 14-day actigraphy, one night polysomnography (PSG) and dim light melatonin onset (DLMO) assessments. Resilience was operationalized as both a coping capacity and a positive mental health outcome in the context of accumulative adversity (life stressful events and childhood adversities). We first identified the covariance patterns between multiple sleep and circadian health domains (Sleep macro- and microstructures, dim light melatonin onset, rest activity pattern) and multifaceted mental resilience using partial least squares regression (PLSR), and followed by univariate analysis as complementary to illustrate the distribution trends of sleep and circadian rhythm parameters across integrative resilience score derived from principal component analysis (PCA).

**Findings:** The PLSR model identified a modest but significant association of greater mental resilience with a sleep and circadian profile (Bootstrapping r = 0.15, 95%CI: 0.02-0.28), characterized by greater frontal-central slow waves activity in NREM sleep, greater frontal theta activity in REM sleep, lower occipital spectral power across fast and slow frequency bands in NREM and REM sleep, and probably earlier dim light melatonin onset timing.

**Conclusions:** Sleep and circadian metrics, specifically frontal slow waves and REM theta activity, attenuated overall occipital spectral power during sleep, and earlier dim light melatonin onset time, represent quantifiable targets that may serve to improve resilience and stress-related mental health in adults. Longitudinal and interventional studies are needed to establish whether sleep interventions can be endorsed as evidence-based approaches for resilience enhancement.

**Clinical implications:** Targeting sleep and circadian aspects may be an important approach to promote stress-related mental health.

**What is already known on the topic:** Sleep and circadian rhythm are theorized to contribute to mental resilience, but the association of an integrated analysis of objective sleep and circadian measures with multifaceted mental resilience remains limited.

**What this study adds:** This study is unique in identifying a modest but significant associations of greater multifaceted mental resilience with a sleep and circadian profile that characterized by greater frontal-central slow waves activity in NREM sleep, greater frontal theta activity in REM sleep, lower occipital power across fast and slow frequency bands in NREM and REM sleep, and probably earlier dim light melatonin onset timing.

**How this study might affect research, practice or policy:** This study shows that sleep and circadian rhythm are linked to resilience. The findings underscore the potential value of interventions designed to modify sleep and circadian domains in order to support stress-related mental health.

## Background

Mental resilience is crucial for health, as it enables individuals to effectively cope with stress, overcome challenges, and maintain emotional well-being[1]. It is a multifactorial concept that encompasses both capacity and outcome of mental health maintenance withstanding a multitude of adversity[1]. While resilient coping is traditionally regarded as a daytime process, emerging studies suggested that there is a diverse neurophysiological process during sleep in conferring mental resilience [2]. For example, slow wave sleep and sleep spindles during Non-rapid Eye Movement (NREM) sleep are suggested to facilitate critical processes such as memory processing, glymphatic clearance, and cortisol regulation, that are essential for stress recovery[3–5]. Additionally, Rapid Eye Movement Sleep (REM) has been postulated to play a crucial role in the mental recovery from stress, possibly via emotional memory processing related to fear extinction and promoting cognitive flexibility that may mitigate stress responses [6,7]. While earlier studies have indicated that sleep microstructures are involved in stress pathophysiology [8–10], recent studies have linked them to positive stress adaptation and resilience[11,12].

Furthermore, a healthy entrained circadian was also theorized to confer resilience[13,14], by modulating the hypothalamus-pituitary-adrenal gland axis, autonomic nervous system, and transcription of genes expression that encodes stress-related hormones, inflammatory markers and neurotransmitters[13,14]. Besides, daily social rhythm cues, such as work schedules and daily routines, introduce daily stressors and play critical roles in regulating circadian rhythms[15,16]. Such synchronization, driven by natural (such as blue light) and social cues, is believed to confer an adaptive advantage by enabling active anticipation of predictable environmental fluctuations, thereby optimizing physiology and behavior over passive reactivity [17]. While morning chronotype was reported to be associated with better mental resilience capacity in healthy and clinical populations[18–22], most studies lack objective circadian measures.

In addition, there remains a lack of standardized definitions of mental resilience[11,12,23]. Some studies defined resilience as the absence of significant psychopathology, mostly referring to the development of post-traumatic stress disorder (PTSD) after a traumatic event[9,24], whereas some studies solely quantified resilience by the level of positive coping tendency[11]. Besides, most studies focused on clinical population and omitted a substantial community population who have remained mentally healthy despite various adversities. Moreover, there is a dearth of study with integrated and objective sleep and circadian rhythm measures that will be correlated with a multidimensional framework of mental resilience. To answer the question, we applied a multivariate data-driven statistical technique to discover the latent dimensions that link interindividual variability of mental resilience to the variability in measures spanning and integrating multiple domains of sleep and circadian rhythm, including macro- and microstructures of sleep, circadian measure by dim light melatonin secretion pattern and rest activity pattern measured by actigraphy.

## Objective

To address the knowledge gap, we aimed to elucidate the association between multifaceted mental resilience and sleep and circadian rhythm metrics in healthy adults who either maintained well-being despite childhood and life adversities or reported no adversity. We hypothesized that the multifaceted mental resilience score would be associated with sleep macro- and microstructures and circadian rhythm. The partial least square regression (PLSR) machine learning model was applied to examine the associations.

## Methods

### Participants

Healthy adults aged between 21 and 45 years with absence of current and lifetime mental disorder, sleep and circadian rhythm disorder were recruited. Exclusion criteria included: 1) have been diagnosed with significant physical disorders, such as epilepsy, brain tumor; 2) on regular medications such as benzodiazepines and antidepressants; 3) pregnancy; 4) shift work schedule; and 5) travel across time zone in the past month. Eligible participants were screened with the clinical version of Structured Clinical Interview for DSM-5 (SCID-CV) and the Diagnostic Interview for Sleep Patterns and Disorders (DISP), for assuring the absence of psychiatric disorder, sleep and circadian rhythm disorder, respectively[25,26]. The study was approved by the Joint Chinese University of Hong Kong-new Territories East Cluster Clinical Research Ethics Committee (No.: 2020.249) and the Institutional Review Board of the University of Hong Kong (EA1909038).

### Measures

All participants were invited to complete online questionnaires and undergo a series of sleep and circadian rhythm assessments. The questionnaire included measures of sociodemographic characteristics, lifestyle (e.g., caffeine and alcohol consumption, and exercise), quality of life, adversities, resilience, sleep and circadian questions. The sleep wake pattern was measured by 14-day actigraphy (Philip Actiwatch Spectrum Pro, Koninklijke Philips N.V.) and concurrent sleep diary. After 7 days record of actigraphy and sleep diary, participants were scheduled with one night polysomnography (PSG), followed by one night dim light melatonin onset (DLMO) assessment, and they received the assessments either in the laboratory or at home. The night of DLMO assessment was not included in actigraphy analysis.

### Resilience Measures

To explore different aspects of resilience, we adopted two complementary but related approaches: one was to define resilience as a capacity trait [23], and the other one was to define resilience as a positive mental health outcome despite previous adversities [27].

### Resilience capacity and resilience outcome measures

Resilience capacity was measured by the Connor-Davidson Resilience Scale (CDRS-10). Resilience outcome was estimated by the standardized residual value of the linear regression model with participants’ average score of quality of life (WHOQOL-BREF) as dependent variable and their cumulative adversities as independent variable. The cumulative adversities were calculated by the sum of participant’s lifetime major stressful events and childhood adversities, measured by the adapted version of Life Stressful Index (LSI) [17], and the Maltreatment and Abuse Chronology of Exposure (MACE)[28], respectively. The adapted LSI [17] assessed fifteen types of stressful events, including natural disasters, serious accidents, exposure to toxic substances, physical assault, sexual assault, unwanted sexual experience, arrested or dispute with in-laws, life-threatening injury, death of family members or close friend, enormous change of relationships among family members and romantic relationships, worsening of financial situations, tremendous workload and significant event in job occasions in the recent 1 year. The MACE scale has 52 items for assessing 10 types of childhood adversities including parental verbal abuse, parental nonverbal emotional abuse, parental physical maltreatment, emotional neglect, physical neglect, witnessing interparental violence, witnessing violence to siblings, sexual abuse, peer emotional abuse, and peer physical bullying[28].

### Integrative resilience measures

Although the resilience trait measure and dynamic adaptation outcome measures are conceptually complementary, they are also moderately correlated and may introduce multicollinearity when being entered simultaneously as predictors in regression models. Moreover, each single indicator alone may contain measurement-specific variance that is orthogonal to the core construct of resilience. To distill the shared variance indicative of the “integrative resilience” phenotype, we conducted a principal component analysis (PCA) on the standardized scores of the two measures. PCA revealed a single component with an eigenvalue of 1.62, which accounted for 81.07% of the total variance. Both scores of CDRS and resilience outcome residual showed high loadings (CDRS: 0.71; Resilience outcome residual: 0.71), indicating that the latent component equally represents trait resources and stress-adaptation outcomes. Therefore, the resulting latent score was labeled as the individual “Integrative resilience score” for each participant, and the participants were grouped into “Low integrative resilience” (<25^th^ percentile), “Moderate integrative resilience” (25-75^th^ percentile) and “High integrative resilience” (>75^th^ percentile).

### Sleep Macro- and Microstructure Measures

The general sleep architecture was measured by video-PSG at the laboratory (Laboratory PSG, sampling rate: 512Hz; Grael PSG, Compumedics Limited) or ambulatory portable PSG at home (Ambulatory PSG, sampling rate: 200 Hz[29]; Nox A1, Nox Medical, Reykjavik, Iceland). Both Laboratory PSG and Ambulatory PSG recording standard montages included electroencephalogram (EEG), bilateral electrooculogram (EOG), electrocardiogram (ECG) and electromyogram (EMG) of mentalis muscle and bilateral anterior tibialis muscles. Bipolar EEG was obtained from frontal (F3, F4), central (C3, C4), and occipital deviations (O1, O2) following the 10-20 systems. The respiratory assessments that included nasal airflow, thoracic and abdominal respiratory efforts and oxygen saturation (SpO2) were also included. A registered PSG technologist (RPSGT) who was blind to the resilience level of participants and two trained researchers manually scored sleep stages in each 30-s epoch according to the American Academy of Sleep Medicine (AASM) 2017 guidelines. The inter-rater reliability of PSG staging ranged between 0.86 and 0.92.

The sleep EEG data was further processed with the sleep analysis toolbox, named “YASA (Yet another Spindle Algorithm)” , implemented using python (Anaconda Jupyter Notebook) [30]. All included EEG signals were down sampled to 100 Hz, and bandpass filtered at 0.1 Hz and 35 Hz. EEG channels including frontal (F3, F4), central (C3, C4), and occipital (O1, O2) were included in calculation. The staged epochs were further tested with automatic artifact rejection based on the distribution of the standard deviations (SD) of each epoch and each EEG channel. To control EEG quality, any epoch with ≥1 channel exceeding 2 SD out of mean value of signal amplitudes would be marked as artifact and excluded. Power spectral density (PSD) was calculated using the Welch method with 4-second sliding windows. The absolute band power value of the following bands was calculated: 1) Slow delta band: 0.1-1 Hz; 2) Fast delta band: 1-3.5 Hz; 3) Theta band: 3.5-8 Hz; 4) Alpha band: 8-12 Hz; 5) Slow sigma band: 12-14 Hz; 6) Fast sigma band: 14-16 Hz; 7) Slow beta band: 16-24 Hz, and 8) Fast beta band: 24-32 Hz. These bands in REM and NREM stages N2 and N3 sleep were calculated for each participant and then log-transformed. We also extracted slow wave sleep and sleep spindles implemented in YASA. In the frontal and central regions, discrete slow waves with a frequency of 0.1-3.5Hz were detected by the adapted algorithms based on previous studies, and sleep spindles were detected with the algorithm that applied threshold of sigma power frequencies of interest (11-16 Hz). Beyond the periodic spectral bands, the aperiodic component of the EEG power spectrum was also extracted, using the fitting algorithm implemented in YASA. For each epoch, the power spectral density in log-log space across the 0.1-32Hz range were extracted, and the spectral slope and the spectral intercept were extracted. The spectral slope indices the excitation-inhibition balance, and the spectral intercept reflects broadband signal amplitude. These parameters were averaged across frontal, central, and occipital electrodes separately for NREM and REM sleep.

### Circadian Measures

Participants were instructed to wear an actiwatch on nondominant wrist and reported sleep-wake timing in a sleep diary for 14 days. Activity data were recorded every 1 minute. The wake threshold was set to medium sensitivity (40 counts per minute), and the time of inactivity for estimating sleep onset and offset timing was set to 10 minutes. The sleep wake time were manually scored according to self-report data from sleep diaries. Actigraphy variables included sleep onset time, sleep onset latency, sleep offset time, wake time after sleep onset, and sleep efficiency. Rest activity pattern was calculated by nonparametric analysis by the “nparACT” package in R studio (2022.07.2). The Interdaily stability (IS), Intradaily variability (IV), Relative amplitude (RA), activity count of the most active 10 hours (M10), active phase onset timing (M10 onset), activity count of the least active 5 hours (L5), and rest phase onset timing (L5 onset) were calculated. Cosinor analysis was also conducted to obtain the acrophase, which reflects the peak timing of the rest-activity pattern.

As for the dim light melatonin onset (DLMO) assessment, the salivary collection period was scheduled starting at 6 hours before individual’s habitual bedtime and ending at 2 hours after habitual bedtime. We collected dim light melatonin salivary samples in both In-laboratory settings and At-home settings (Details were reported in supplementary information). Salivary melatonin samples were analyzed by the Liquid Chromatography-Mass Spectrometry (LCMS) method, using the positive electrospray ionization (Waters Acquity Xevo TQ-XS system, Waters Corporation, Milford, MA, USA). DLMO time was determined when the concentration of melatonin reached the threshold of 3pg/mL and kept above the threshold the next two hours.

### Statistical analysis

All statistical analyses were performed using RStudio (2022.07.2) and Python 3.9 within the Anaconda distribution (Jupyter Notebook), and the significance level was set at two-tailed p <0.05. Given the collinearity characteristics of sleep and circadian rhythm variables, we first used partial least squares regression (PLSR) model to predict mental resilience from sleep and circadian rhythm variables. To contextualize the multivariate PLSR findings, we performed further univariate analysis as descriptive summaries.

### Step 1: Partial Least Squares Regression (PLSR)

We applied PLSR, a machine-learning technique that seeks to find covariance between multidimensional matrices. Given the large number of features (absolute power in different frequency bands across frontal, central and occipital region across different sleep stages, sleep macrostructures, and circadian phase variables; k=210), and the sample size (n=156), and the high level of correlation among the features, we applied the PLSR model to characterize the association between sleep and circadian rhythm and mental resilience. Herein, the outcome matrix *Y* consisted of two mental resilience indicators: resilience capacity, indexed by the CDRS score, and resilience outcome, indexed by the residual from the linear regression model predicting quality of life from accumulative adversities. The higher residual value indicated better-than-expected quality of life given the level of accumulative adversity and were therefore interpreted as better resilience outcomes.

The predictors *X* included sleep macro- and microstructures and circadian phase features. We applied feature selection to sleep macro- and microstructural variables separately for NREM and REM sleep, because there is a large number of candidate features regarding power spectral density, aperiodic activity, slow waves and sleep spindles characteristics across electrodes[31]. In contrast, circadian rhythm variables consisted of aggregated measures (e.g., interdaily stability, intradaily variability, DLMO time) and were therefore retained and entered into the PLSR model together with the selected NREM and REM macro- and microstructural features. We used the fully nested cross-validation framework for isolating feature selection from model evaluation[32]. To avoid data leakage, standardization of the predictors *X (sleep macro- and microstructures)* and outcome matrix *Y* were conducted within the training set, and all predictor selection procedures were embedded within the training loop of the cross-validation. For each outer training set, feature selection was performed using an inner 5-fold stability procedure. Within each inner training split, the feature was retained as a predictor in the PLSR model if the p-value of the univariate correlation was significant (p<0.05). Feature selection frequency across the 5 inner folds was then evaluated using the binomial test assuming a null selection probability of 0.05, and features with binomial p<0.05 was retained, given the threshold p-value selected. Only features with selection ratio > 80% were retained as stable predictors for the current outer training set.

The selected NREM and REM sleep macro- and microstructural features were further applied for PLSR model training, together with circadian features that included DLMO time, interdaily stability, interdaily variability, relative amplitude, activity on active and rest period (M10 and L5), onset time of rest period and active period, and acrophase. To determine the optimal number of latent components, we performed a grid search over *n_components* ranging from 1 to 10 using cross-validation in each inner fold. For each candidate number, we calculated the Pearson correlation coefficient and RMSE between observed and predicted values in each left-out fold. The optimal number was selected by prioritizing the minimization of mean RMSE, while considering the stability of correlation coefficients to balance goodness-of-fit and generalizability. One component was selected in most outer folds and was therefore used for interpretation (Component = 1, mean r = 0.22, mean RMSE = 3.35). We then used the 10-fold cross validation to estimate out-of-sample predictive performance. We calculated the Pearson r between predicted and observed values for resilience capacity score (CDRS) and resilience outcome (residual value) separately, and then averaged them to represent the overall model effect size (mean r). To estimate the sampling variability of the mean test-set correlation, we performed 10,000 bootstrap resamples of the participants (with replacement) on the paired observed and out-of-sample predicted values. The analysis was conducted on the fixed predictions from the outer CV folds and did not involve re-fitting models in each bootstrap iteration. We further adjust for covariates within each training fold by residualizing both predictors and outcomes with respect to age, sex, educational level and family income.

### Step 2: Univariate analysis

We performed further univariate analysis of sleep macro- and microstructures, and circadian rhythm variables. We derived the integrative resilience latent score from principal component analysis (PCA) based on the score of resilience capacity and resilience outcome, with higher scores indicating greater multidimensional resilience. Correlational analyses were conducted between the score of integrative resilience and sleep macro- and microstructures and circadian rhythm parameters. In addition, the descriptive statistics of sleep macro- and microstructures and circadian rhythm parameters across integrative resilience groups (using 25^th^ and 75^th^ cutoff) were presented. Notably, as these univariate analyses were for descriptive purpose and not intended for hypothesis testing, the significance were not interpreted.

## Findings

### Participants

The total sample consisted of 156 participants (Mean age: 29.54 years, Range: 21-48 years, 60.9% female; Table 1). Among them, 37 were identified as with low integrative resilience, 80 as with moderate integrative resilience and 39 as with high integrative resilience. Participants with higher-level integrative resilience showed higher-level educational background, lower sleep reactivity and less mental distress, and probably more morning chronotype (Table 1).

**Table 1.** Demographics, sleep and mental health characteristics across resilience groups.

|  | All sample | Low integrative resilience | Moderate integrative resilience | High integrative resilience | p value |
| --- | --- | --- | --- | --- | --- |
| **Sample size** | **156** | **37** | **80** | **39** |  |
| **Age**, years, Mean±SD | 29.54±4.95 | 29.54±5.28 | 29.20±4.33 | 30.26±5.83 | 0.55 |
| **Sex**, n (%) |  |  |  |  |  |
| Female | 95 (60.9) | 27 (73.0) | 47 (58.8) | 21 (53.8) | 0.20 |
| Male | 61 (39.1) | 10 (27.0) | 33 (41.3) | 18 (46.2) |  |
| **Educational level**, n (%) |  |  |  |  |  |
| ≤ High school | 5 (3.2) | 4 (10.8) | 1 (1.3) | 0 | 0.01 |
| ≥ Bachelor's degree | 151 (96.8) | 33 (89.2) | 79 (98.8) | 39 (100.0) |  |
| **Marital status** |  |  |  |  |  |
| Cohabit | 4 (2.6) | 1 (2.7) | 2 (2.5) | 1 (2.6) | 0.84 |
| Married | 34 (21.8) | 8 (21.6) | 16 (20.0) | 10 (25.6) |  |
| Divorced | 2 (1.3) | 0 | 2 (2.5) | 0 |  |
| Widowed | 2 (1.3) | 0 | 2 (2.5) | 0 |  |
| Single | 114 (73.1) | 28 (75.7) | 58 (72.5) | 28 (71.8) |  |
| **Employment status**, employed, n (%) | 106 (67.9) | 26 (70.3) | 55 (68.8) | 25 (64.1) | 0.83 |
| **Chronotype**, n (%) |  |  |  |  |  |
| Morning type | 26 (16.7) | 6 (16.2) | 11 (13.8) | 9 (23.1) | 0.07 |
| Intermediate type | 91 (58.3) | 19 (51.4) | 45 (56.3) | 27 (69.2) |  |
| Evening type | 39 (25.0) | 12 (32.4) | 24 (30.0) | 3 (7.7) |  |
| **Sleep vulnerability**, n (%) |  |  |  |  |  |
| High sleep reactivity | 90 (57.7) | 31 (83.8) | 45 (56.3) | 14 (35.9) | <0.001 |
| **Mental distress (GHQ-28)** |  |  |  |  |  |
| Total score | 17.35±9.52 | 22.00±11.69 | 18.39±8.61 | 10.79±4.35 | <0.001 |
| Somatic | 4.69±3.37 | 5.43±3.86 | 5.18±3.23 | 3.00±2.52 | 0.001 |
| Anxiety | 4.17±4.03 | 5.43±4.75 | 4.84±3.82 | 1.59±2.29 | <0.001 |
| Depression | 1.67±2.77 | 2.97±3.46 | 1.74±2.73 | 0.28±0.94 | <0.001 |
| Social dysfunction | 6.82±2.44 | 8.16±2.41 | 6.64±2.22 | 5.92±2.43 | <0.001 |
| **Quality of life (WHOQOL-BREF)** |  |  |  |  |  |
| Average score | 65.67±11.74 | 52.76±8.48 | 64.81±6.45 | 79.67±6.55 | <0.001 |
| Psychological health | 62.48±15.20 | 46.84±10.54 | 61.34±10.46 | 79.67±8.08 | <0.001 |
| Physical health | 72.76±12.58 | 60.86±8.62 | 71.66±9.58 | 86.31±7.48 | <0.001 |
| Social relationship | 62.86±15.62 | 51.35±16.31 | 62.08±12.15 | 75.38±12.02 | <0.001 |
| Environment | 64.56±14.18 | 52.00±12.84 | 64.15±10.13 | 77.31±11.39 | <0.001 |

Note: Data in the table are presented as n (%) or mean±SD. Differences in demographics, sleep and mental health characteristics across resilience groups were performed with analysis of variance (ANOVA) test. Categorical variables were compared by Chi-square test or Fisher's exact test. Results of post-hoc Bonferroni corrections showed that the levels of mental distress decrease across integrative resilience (Low integrative resilience > Moderate integrative resilience > High integrative resilience, all p values < 0.05), while the level of quality of life increase across integrative resilience groups (Low integrative resilience < Moderate integrative resilience < High integrative resilience, all p values < 0.05)

### PLSR model identified a dimension linking slow wave sleep, REM sleep theta power, and probably early circadian phase and mental resilience

After feature selection within the inner loops of the nested cross-validation procedure, 30 of 145 NREM sleep predictors and 5 of 56 REM sleep predictors with selection ratio exceeding 0.50 was selected for analysis. The PLSR model was fitted using both the selected sleep macro- and microstructural features and the circadian rhythm parameters as independent variables. Figure 2A shows the Variable Importance in Projection (VIP) scores of features, and slow wave sleep (delta power, negative peak amplitude, density) in NREM sleep, frontal theta and fast delta power (1-8Hz) and occipital delta power in REM sleep, and the percentage of REM sleep had VIP scores exceeding 1. The DLMO time ranked relatively less high (VIP≈0.8), suggesting a weaker but potentially relevant contribution. Figure 2B further demonstrates the relative importance of contributors by showing the weights and loadings. The results of loadings and weights showed that greater frontal slow waves activity (greater delta power, and greater density), greater frontal theta power in REM sleep, and less occipital delta and theta power in NREM sleep and REM sleep were associated with higher composite score of resilience. DLMO time had relatively small negative weight and loading values (weight = -0.13, loading = -0.05), indicating that earlier DLMO was marginally associated with higher resilience. The Bootstrap correlation coefficient was 0.15, with 95% confidence interval ranging between 0.02 and 0.28 (Figure 2D). We further adjusted for age, sex, educational level and family income in each training fold, and the results remained largely unchanged (Supplementary Figure S1; Bootstrap r = 0.20, 95%CI: 0.06-0.34). Collectively, these multivariate patterns indicated the association of higher mental resilience with an integrated sleep–circadian profile characterized by higher frontal slow-wave sleep activity, higher frontal REM theta activity, lower occipital slow and fast frequency band power in NREM and REM sleep, and probably an earlier circadian phase.

**Figure 1.**
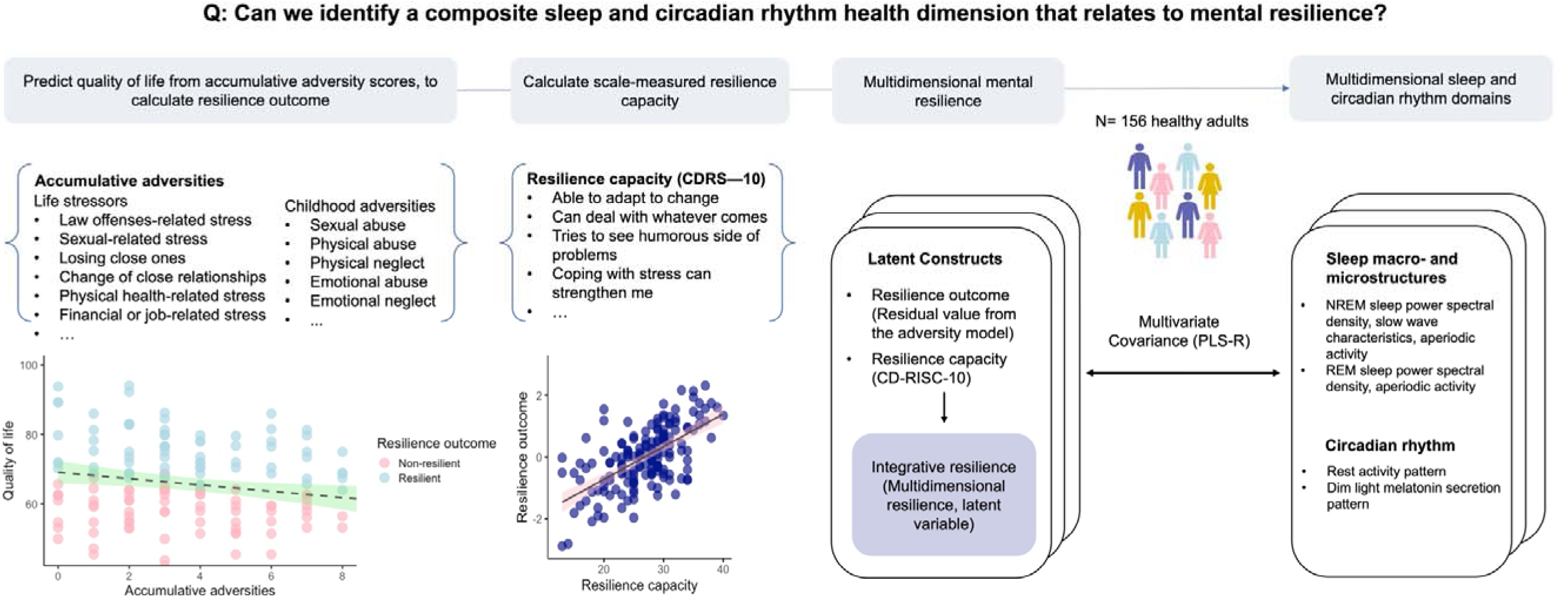
Study design diagram, from integrative resilience calculation to integration of sleep and circadian rhythm variables and their association with integrative resilience.

**Figure 2.**
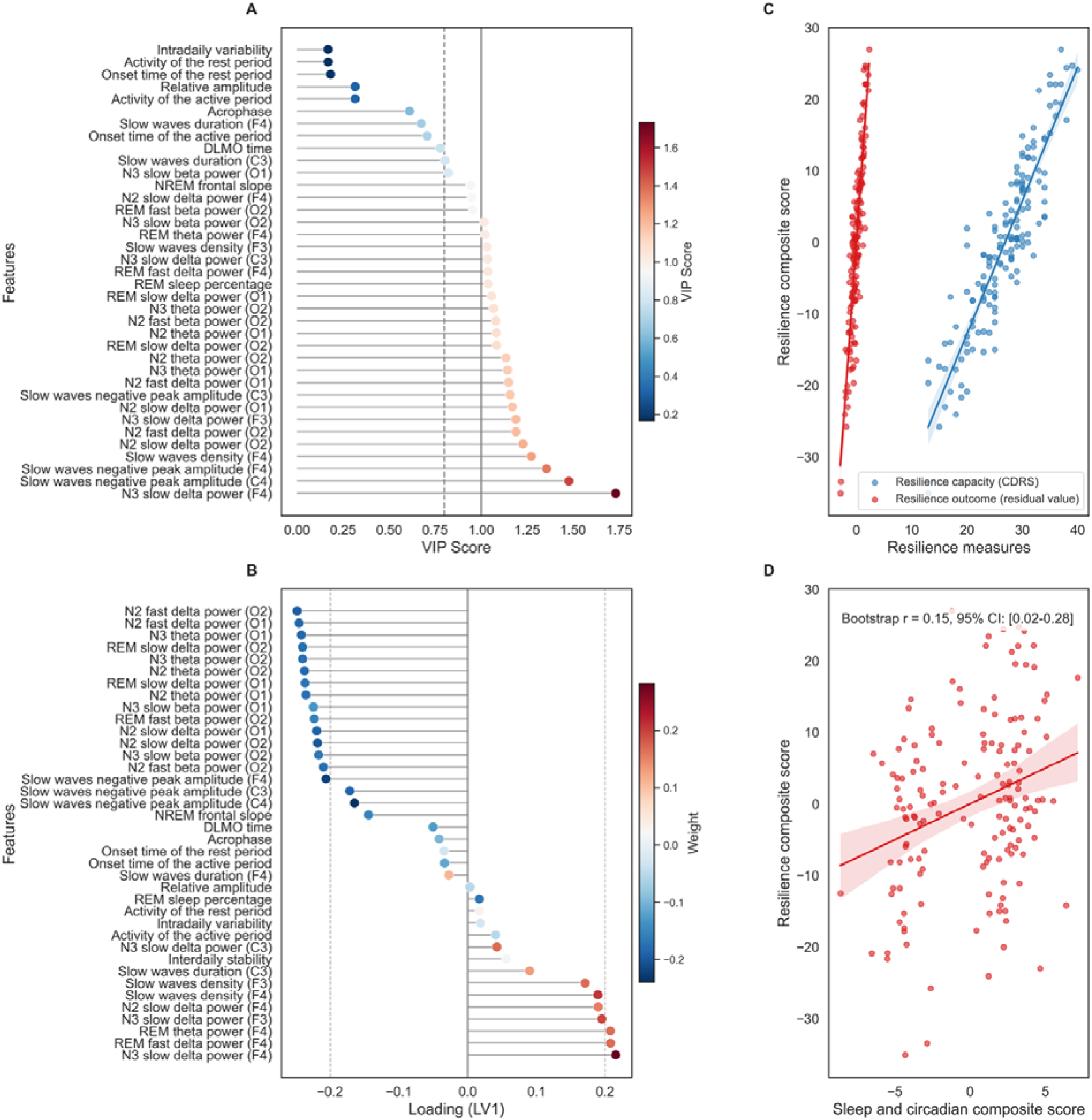
PLSR reveals one dimension linking sleep and circadian health and mental resilience. Note: A. VIP scores of final selected predictors. The color gradient (blue to red) indicates increasing predictive importance, with the dashed gray line denoting the conventional VIP=0.8, and the solid line denoting VIP=1.0 benchmark. Higher VIP scores reflect stronger contribution to the multivariate covariance structure between the features and mental resilience. B. Weight and Loading scores of final selected predictors. Bar colors represent the standardized weights of each predictor for LV1 (red: positive contribution; blue: negative contribution). The slow wave characteristics that include higher slow delta power, greater slow waves density, and greater negative peak amplitude of slow waves significantly contribute to LV1, while earlier DLMO time had marginal contribution. C. Correlation between the PLSR-derived resilience composite score and resilience original scores. D. Scatter plot showing the correlation between the first latent variable derived from the sleep and circadian features (X-LV1, representing the sleep/circadian composite score) and the first latent variable derived from the resilience measures (Y-LV1, representing the resilience composite score). The solid red line represents the linear fit, with the shaded band indicating the 95% confidence interval (Bootstrap r = 0.15, 95%CI: 0.02-0.28).

### Univariate analyses of sleep macro- and microstructures, and circadian rhythm with mental resilience

As shown in Figure 3A, the higher integrative resilience was correlated with lower REM sleep percentage (Spearman’s rho = -0.17) and longer NREM stage 3 sleep duration (Spearman’s rho = 0.17). Descriptive statistics of sleep macrostructures across integrative resilience groups were detailed in Supplementary Table S1. Regarding sleep microstructures, higher integrative resilience was associated with increased slow-delta power over frontal and central regions during NREM stage 3 sleep, with the association being relatively more prominent over the right frontal region (Figure 3B; Detailed in Supplementary Table S2-S4). In occipital regions, higher integrative resilience was associated with lower power across the slow delta, fast delta and theta bands (0.1-8Hz) during NREM stage 2 sleep, as well as reduced power in the slow delta (0.1-1Hz) and fast beta (24-32Hz) bands during REM sleep. Descriptive comparisons of sleep microstructures across the three integrative resilience groups are presented in Supplementary Figure S2 and Supplementary Table S5-S8.

**Figure 3.**
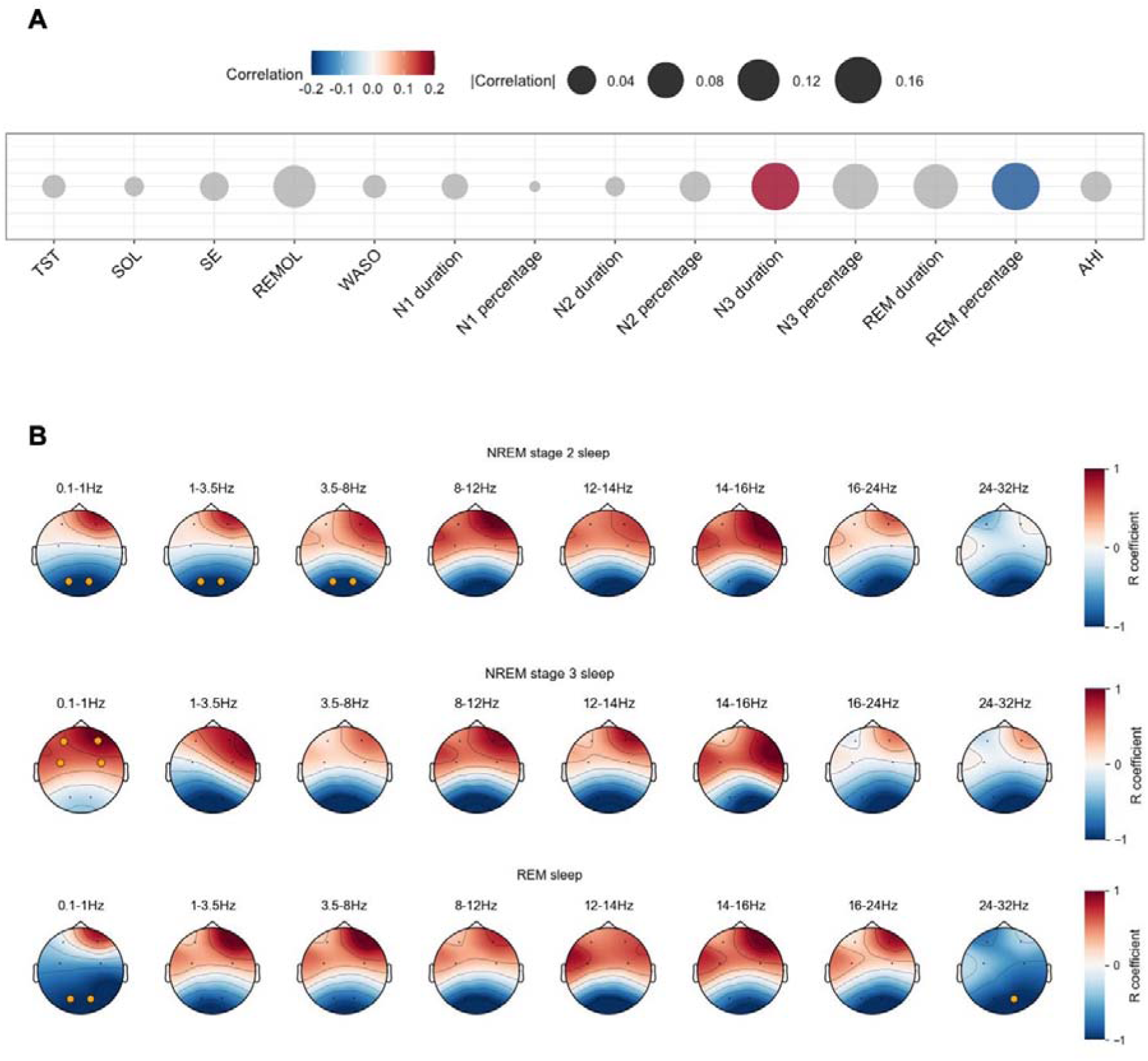
Univariate correlational analysis examining associations between integrative resilience (latent score) and sleep macro- and microstructures. Note: A. Bubble plot showing correlations between integrative resilience (latent score) and sleep macrostructures. A longer duration of NREM stage 3 sleep and greater percentage of REM sleep was correlated to higher latent score of mental resilience (significance shown in colored bubbles). B. Topoplots showing correlations between log-transformed absolute power in frequency bands in NREM and REM sleep, and latent score of mental resilience. The yellow dotted points showed electrodes with significance (Pearson’s correlation, p < 0.05).

For circadian rhythm features (shown in Figure 4), an earlier dim light melatonin onset (DLMO) time was associated with a higher integrative resilience score (Spearman’s rho = -0.20). The median DLMO times were 22:56, 22:43 and 22:19 for the low, moderate, and high integrative resilience groups, respectively. Throughout the sampling period, the high integrative resilience group showed a high overall melatonin secretion level trajectory (Figure 4A). The timepoint specific descriptive comparisons of melatonin level during sample period were provided in Supplementary Tables S9. No associations were found between actigraphy-derived sleep wake pattern and rest activity pattern metrics and multifaceted mental resilience. The correlation coefficients of integrative resilience with DLMO time and rest activity pattern were detailed in Supplementary Table S10.

**Figure 4.**
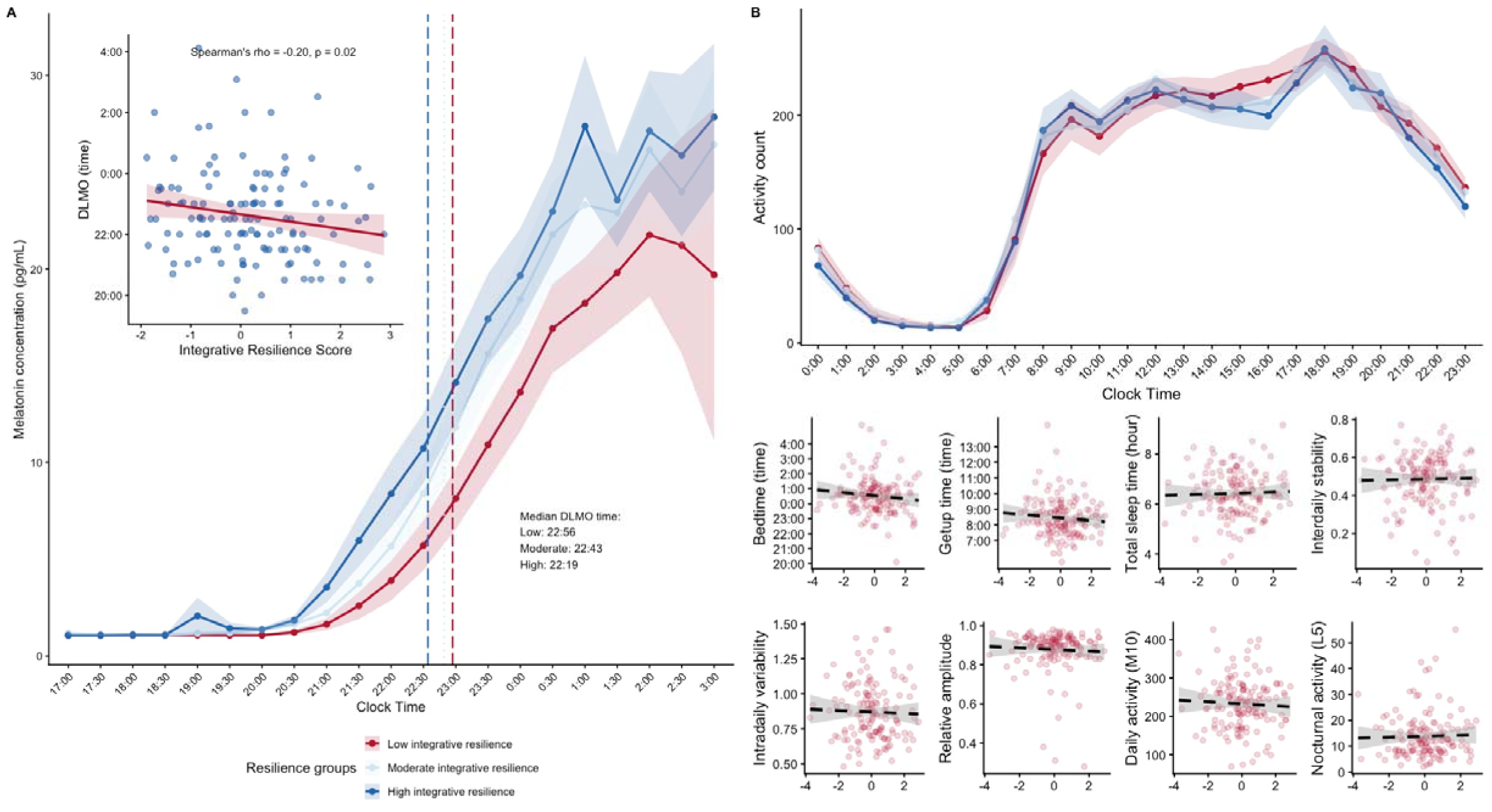
Integrative resilience and circadian rhythm. Note: A. Dim light melatonin secretion pattern across time from 5pm to 3am in this study, with embedded scatter plot showing the correlation between DLMO timing and integrative resilience score (Spearman’s rho = -0.2, p=0.02), suggesting that earlier circadian phase was associated with higher resilience. B. The 24-hour activity pattern across integrative resilience groups, and scatter plots showing the association between integrative resilience score and rest activity pattern and sleep wake pattern, as measured by the 14-day actigraphy.

## Discussion

This hypothesis-generating and exploratory study examined the correlates of sleep and circadian measures with an integrated multidimensional concept of mental resilience by taking into account of both capacity and outcome measures in healthy adults. Our findings revealed a modest coherent sleep and circadian profile of mental resilience, characterized by enhanced frontal slow-wave activity, and frontal theta power in REM sleep, suppressed occipital fast and slow frequency power in both NREM and REM sleep, and a tendency toward earlier circadian phase. Although the cross-validated predictive effect was modest in magnitude, it survived bootstrap resampling, suggesting that the collective contribution of sleep and circadian factors to mental resilience may be valid and meaningful.

Importantly, slow wave activity was of great value to mental resilience, which was consistent across both PLSR and univariate analyses. Slow delta power over frontal region and slow waves negative peak amplitude over frontal-central region during NREM stage 3 sleep emerged as strong contributors to the latent component of resilience capacity and resilience outcome. Complementary to the frontal slow wave characteristics, reduced power in slow and fast frequency bands over the occipital region in NREM and REM sleep were also associated with higher resilience, suggesting a topographically distinct cortical signature of resilience, potentially characterized by frontal-occipital gradient EEG power engagement during sleep [33]. Frontal-central slow waves are hypothesized to reflect synaptic downscaling, a restorative process that renormalizes synaptic strength following waking experiences and conserves neural responses for adaptive functioning[34]. Individuals with greater slow wave sleep may therefore possess enhanced capacity for emotional regulation and cognitive flexibility, which are important for maintaining resilience[35]. While prolonging NREM sleep has been reported to promote resilience against social defeat stress paradigm in mice [36], there was a lack of similar evidence in human resilience studies, probably because of the complexity of resilience definition, or that most studies predominantly focused on pathological perspectives of sleep after stress exposure [1]. Nonetheless, an experimental study of playing sound cues of positive words during NREM sleep would enable EEG theta power and slow oscillation, which ameliorated the aversive content of memories of emotional pictures [37]. The experimental findings was concordance with our data that slow wave sleep serves as a protective factor for resilience. Our finding is also in line with previous findings of widespread recovery function in slow wave sleep, such as inhibiting hypothalamus pituitary adrenal (HPA) secretion of cortisol[38], enhancing neural activity and cognitive processing[12,39], and driving metabolic glymphatic clearance from the brain[40]. Thus, the slow wave sleep activity in NREM sleep, may facilitate widespread physiological stress recovery functions, therefore conferring mental resilience.

The REM sleep findings, particularly the association between elevated right frontal theta power and higher resilience, extend the current understanding of REM sleep’s role in emotional processing. REM theta oscillations, typically within the 3.5-8Hz range, have been implicated in the offline facilitation of emotional memory[6], potentially enabling individuals to reappraise negative experiences and maintain mental resilience. Interestingly, while greater frontal theta power in REM sleep was associated with higher resilience, a greater REM sleep percentage was intriguingly associated with lower resilience. Previous studies indicated that REM sleep length may reflect the neuro-state of emotional activity[41,42], and abnormal REM sleep quantity after stress and adversity might be pathological and maladaptation to stress[41,42]. For example, a prior study found that the longer REM sleep was associated with overnight dissipation in amygdala reactivity, suggesting detrimental function of longer REM sleep for mood processing[43]. Together, these findings prompt us to consider that, excessive REM sleep might constitute a risk factor for future stress problems that demand attention[42,44,45], whereas an appropriate and optimal amount of REM sleep and enhanced frontal theta power may support emotional memory processing and contribute to resilience. Further research is needed, particularly with larger sample size and experimental design, to clarify the associations between REM sleep macrostructure, REM-related theta oscillations, and resilience.

In addition, our findings also suggest a modest potential contribution of early DLMO timing to higher resilience. Prior work demonstrated links between morning chronotype (circadian preference) and improvement in mental well-being, and decreased risk of mental disorder[19–21]. Personality traits like conscientiousness and openness[46,47], and adaptive coping like positive reframing and optimism [48,49], which were considered facilitating individuals in adapting stress, have also been reported as salient characteristics in people with early circadian preference. However, the weight and loading of DLMO time in the PLSR model were lower than those of NREM and REM sleep features. We speculate that it likely captures shared variance with other behavioral factors, such as morning-oriented personality traits and adaptive coping styles, rather than representing a direct neurobiological substrate of resilience. Therefore, the early DLMO timing may be seen as a secondary supportive factor of resilience. Future studies are needed to clarify such speculation.

It could be argued that resilience and sleep may covary from day to day because stress exposure and stress adaptation fluctuate[50]. However, the latent components linking sleep and circadian measures to multifaceted resilience were driven primarily by sleep EEG microstructure and possibly DLMO timing, rather than by 14-day actigraphy-derived rest-activity rhythms. The findings in this study may suggest that the multifaceted mental resilience encompassing positive coping capacity and positive mental health outcome despite accumulative adversities, may be more driven by sleep microstructures and DLMO that capture neurophysiological sleep and circadian processes than by behavioral rest activity patterns shaped by social demands, light exposure, and lifestyle factors. However, these findings remain exploratory, and future studies combining multi-night PSG with ecological momentary assessment are needed to clarify these possibilities.

To the best of our knowledge, this is the first study with multi-domain assessment of subjective and objective measurements of macro- and microsleep characteristics, actigraphy-derived rest activity pattern, and melatonin-based biological circadian phase and mental resilience in a comprehensive manner. While earlier studies have suggested the association of resilience with morning chronotype and better sleep quality using questionnaire and polysomnography assessments[11,19], the current study provided a comprehensive series of integrated sleep and circadian measures with polysomnography, actigraphy, DLMO, questionnaire and clinical assessment in healthy subjects. In the absence of confounding influence of psychiatric, physical and sleep disorders, this study was able to tease out the role of sleep and circadian measures in mental resilience more precisely. Moreover, our study addressed the knowledge gaps regarding sleep macro- and microstructures, circadian rhythm profiles and an integrated concept of resilience. The findings will pave the way to develop sleep and circadian intervention to promote mental resilience in the prevention and early intervention of stress psychopathology in vulnerable populations.

However, several limitations should be noted. First, the inclusion of only healthy subjects constituted both strength and limitation. The implications of our results could not be directly inferred to clinical populations, notably stress-related disorders (e.g., PTSD, depression, anxiety disorder), sleep and circadian disorders (e.g., insomnia disorder, obstructive sleep apnea, delayed circadian phase disorder). Secondly, our cross-sectional observational and correlational evidence limited any causal inferences. Longitudinal and experimental studies are needed to clarify the causality among N3 sleep delta power, REM sleep theta power, circadian timing, and mental resilience. Finally, the current methodology which employed the capacity and outcome concept of resilience might only provide insight into the chronic stress spectrum. Further study with acute and chronic stress exposure paradigm may need to further explore sleep and circadian factors in conferring mental resilience.

## Clinical implications

In summary, our study provided evidence that an integrated sleep and circadian phenotype, defined by increased frontal-central slow waves activity in NREM sleep, increased frontal REM theta power, lower occipital spectral power across fast and slow frequency bands in NREM and REM sleep, and probably earlier circadian phase, is associated with higher mental resilience. Our findings support the need for further experimental and interventional studies to determine causality and promote resilience development.

## Data Availability

All data produced in the present study are available upon reasonable request to the authors.

## Acknowledgement

We thank all participants for their cooperation in this study. We also thank the Department of Chemical Pathology, Faculty of Medicine, The Chinese University of Hong Kong, for assistance in analyses of salivary melatonin. This study was funded by the Hong Kong Research Grant Council Collaborative Research Fund (C7069-19GF; PI: Prof Tatia Mei-Chun Lee). The funder has no role in the design and conduct of this study.

## Data availability statement

The data that support the findings of this study are available on reasonable request from the corresponding authors (YKW, SXL and TMCL). The data are not publicly available due to ethical restrictions.

