## Supplementary Figure S1 for "Exploring multifaceted mental resilience from the perspectives of sleep and circadian rhythm in healthy adults"

Chris Xie Chen *et al.*

Tatia Mei Chun Lee,

Shirley Xin Li,

**Supplementary Figure S1. Sensitivity analysis in PLSR identified the modest dimension linking sleep and circadian health and mental resilience, after adjusting for age, sex, educational level and family income.**

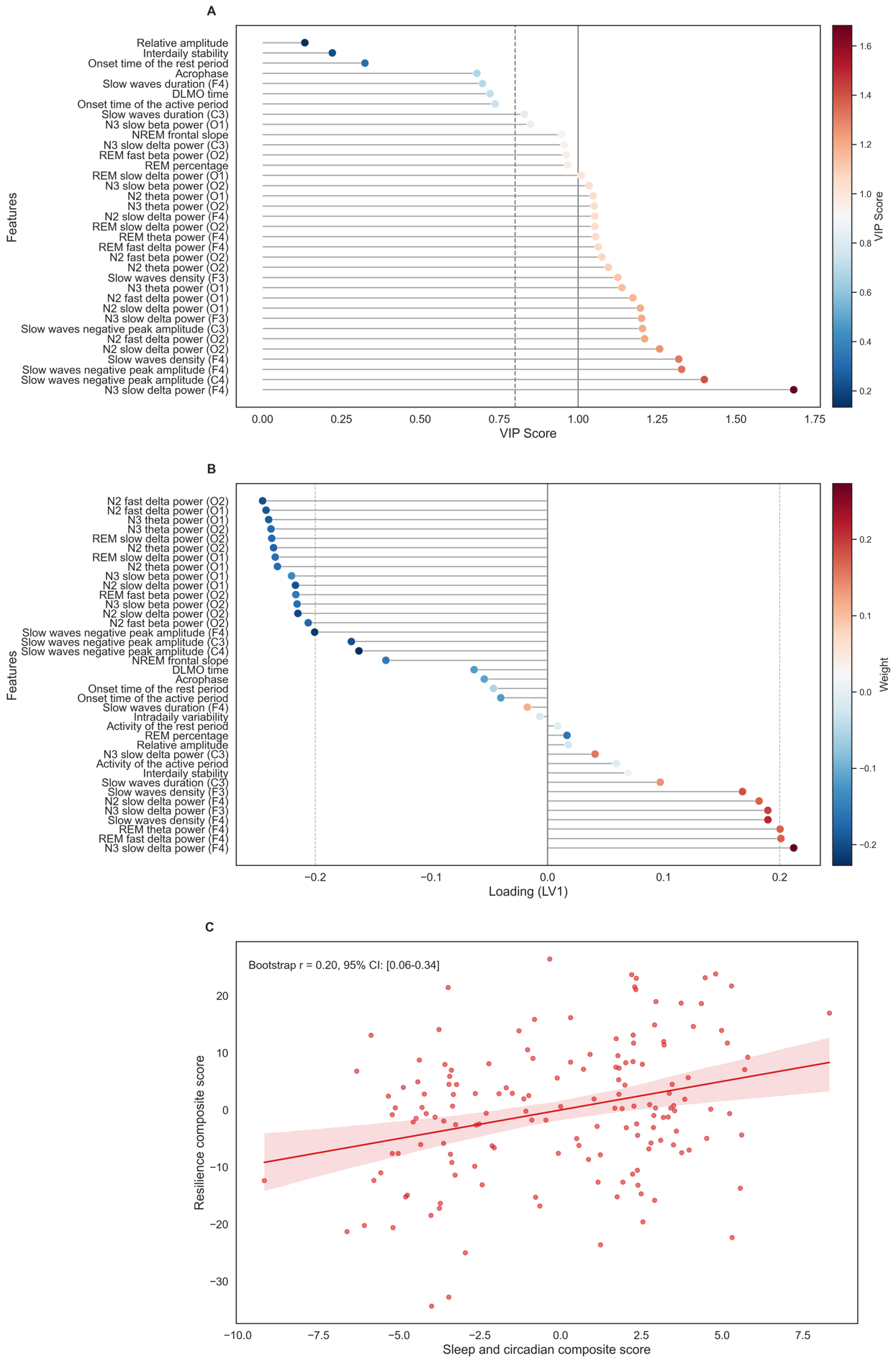

Note: A. VIP scores of selected predictors after covariate adjustment. B. LV1 loadings and weights for individual features. C. Scatter plot of sleep and circadian composite scores and resilience composite scores, after adjustment of age, sex, educational level and family income, via nested cross-validation. Bootstrap resampling (10,000 iterations) found a significant correlation of r = 0.20 (95%CI: 0.06-0.34).

**Supplementary Figure S2. Integrative resilience and sleep microstructures.**

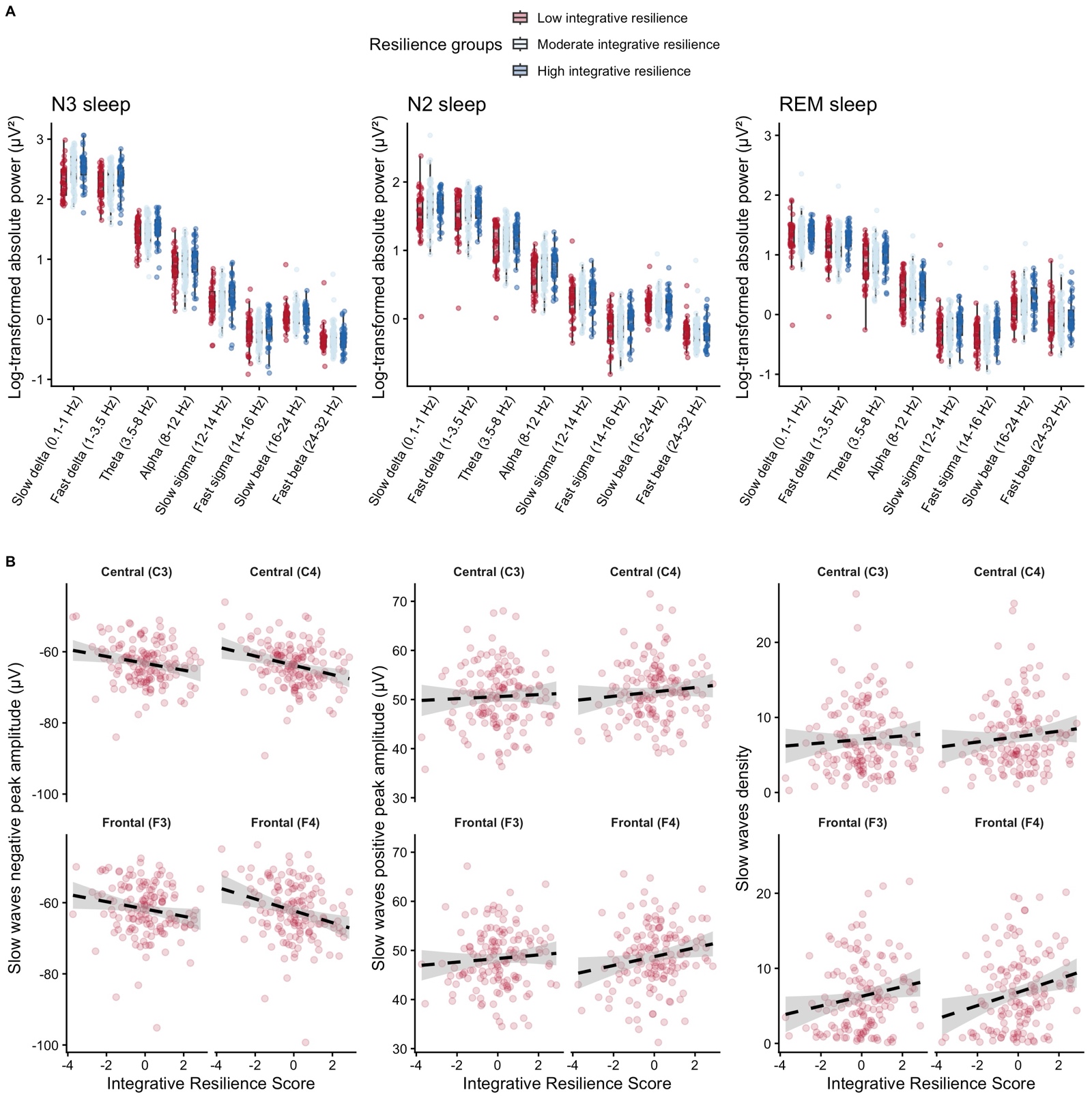

Note: A. Comparisons of average log-transformed absolute power (µV²) of frequency bands in NREM stage 2, stage 3 sleep and REM sleep across integrative resilience groups. B. Scatter plots showing the correlations between integrative resilience score and negative peak amplitude of slow waves in frontal and central regions.

**Supplementary Table S1. Descriptive statistics of sleep macrostructures across integrative resilience groups.**

| **Variable** | **Low integrative resilience** | **Moderate integrative resilience** | **High integrative resilience** |
| --- | --- | --- | --- |
| Total sleep time (min) | 426.81 (16.07) | 423.16 (7.05) | 420.05 (12.34) |
| Sleep onset latency (min) | 11.09 (1.25) | 13.82 (2.02) | 13.53 (1.65) |
| Sleep efficiency (%) | 89.37 (1.82) | 89.39 (0.81) | 88.78 (1.17) |
| Wake time after sleep onset (min) | 39.18 (7.75) | 36.74 (3.59) | 39.44 (5.47) |
| NREM stage 1 sleep (%) | 6.92 (0.97) | 6.27 (0.48) | 5.80 (0.58) |
| NREM stage 1 sleep duration (min) | 29.66 (4.83) | 26.10 (1.92) | 24.76 (2.29) |
| NREM stage 2 sleep (%) | 52.80 (1.35) | 58.85 (5.94) | 55.26 (1.12) |
| NREM stage 2 sleep duration (min) | 225.49 (9.54) | 223.17 (5.19) | 231.80 (8.48) |
| NREM stage 3 sleep (%) | 14.14 (1.11) | 15.13 (0.81) | 15.67 (0.92) |
| NREM stage 3 sleep duration (min) | 58.32 (4.10) | 63.35 (3.49) | 64.90 (3.46) |
| REM sleep (%) | 26.15 (1.16) | 25.75 (0.76) | 23.08 (1.04) |
| REM sleep onset latency (min) | 109.63 (10.61) | 112.77 (6.47) | 130.25 (12.55) |
| REM sleep duration (min) | 113.61 (7.17) | 109.69 (4.03) | 98.32 (5.8) |
| Apnea-hypopnea index (AHI) | 0.86 (0.24) | 0.66 (0.12) | 0.69 (0.19) |

Note: Normally distributed parameters are summarized as mean (SD), whereas non‑normal ones are reported as median (interquartile range).

**Supplementary Table S2. Correlations of integrative resilience (latent score) with NREM stage 2 sleep microstructures.**

| **Feature** | **r coefficient** | **p value** | **Block** |
| --- | --- | --- | --- |
| N2 slow delta power (O1) | -0.19 | 0.02 | N2 |
| N2 fast delta power (O1) | -0.19 | 0.02 | N2 |
| N2 theta power (O1) | -0.18 | 0.03 | N2 |
| N2 slow delta power (O2) | -0.20 | 0.01 | N2 |
| N2 fast delta power (O2) | -0.20 | 0.01 | N2 |
| N2 theta power (O2) | -0.19 | 0.02 | N2 |
| N2 fast beta power (O2) | -0.18 | 0.03 | N2 |
| N2 slow beta power (O2) | -0.17 | 0.04 | N2 |
| N2 fast beta power (O1) | -0.14 | 0.08 | N2 |
| N2 slow delta power (F4) | 0.16 | 0.05 | N2 |
| N2 fast delta power (F4) | 0.14 | 0.08 | N2 |
| N2 theta power (F4) | 0.14 | 0.07 | N2 |
| N2 alpha power (F4) | 0.14 | 0.08 | N2 |
| NREM frontal aperiodic slope | -0.16 | 0.06 | N2 |
| NREM occipital aperiodic intercept | -0.15 | 0.07 | N2 |
| N2 alpha power (O2) | -0.14 | 0.09 | N2 |
| N2 slow sigma power (O2) | -0.13 | 0.09 | N2 |
| NREM frontal aperiodic intercept | 0.14 | 0.10 | N2 |
| Spindles amplitude (F4) | 0.13 | 0.11 | N2 |
| N2 alpha power (O1) | -0.12 | 0.13 | N2 |
| N2 slow beta power (O1) | -0.13 | 0.12 | N2 |
| N2 fast sigma power (F4) | 0.12 | 0.13 | N2 |
| Spindles absolute power (F4) | 0.13 | 0.11 | N2 |
| N2 slow sigma power (O1) | -0.12 | 0.15 | N2 |
| N2 fast sigma power (O2) | -0.11 | 0.17 | N2 |
| NREM central aperiodic intercept | 0.10 | 0.21 | N2 |
| N2 slow sigma power (F4) | 0.09 | 0.26 | N2 |
| N2 fast sigma power (C4) | 0.09 | 0.27 | N2 |
| N2 slow beta power (F4) | 0.09 | 0.26 | N2 |
| Spindles amplitude (F3) | -0.10 | 0.24 | N2 |
| N2 alpha power (C4) | 0.09 | 0.29 | N2 |
| N2 alpha power (F3) | 0.08 | 0.32 | N2 |
| N2 alpha power (C3) | 0.07 | 0.35 | N2 |
| N2 fast sigma power (C3) | 0.07 | 0.35 | N2 |
| N2 fast sigma power (O1) | -0.07 | 0.36 | N2 |
| N2 fast beta power (F3) | -0.07 | 0.35 | N2 |
| Spindles absolute power (C4) | 0.08 | 0.31 | N2 |
| N2 slow sigma power (F3) | 0.06 | 0.43 | N2 |
| N2 theta power (C4) | 0.07 | 0.41 | N2 |
| N2 slow sigma power (C4) | 0.06 | 0.44 | N2 |
| Spindles amplitude (C4) | 0.07 | 0.40 | N2 |
| Spindles duration (F3) | -0.06 | 0.44 | N2 |
| NREM central aperiodic slope | -0.07 | 0.40 | N2 |
| N2 fast sigma power (F3) | 0.06 | 0.45 | N2 |
| NREM occipital aperiodic slope | 0.06 | 0.46 | N2 |
| Spindles duration (C4) | -0.06 | 0.48 | N2 |
| N2 slow sigma power (C3) | 0.06 | 0.49 | N2 |
| Spindles duration (C3) | -0.05 | 0.51 | N2 |
| Spindles duration (F4) | -0.05 | 0.54 | N2 |
| N2 theta power (C3) | 0.05 | 0.56 | N2 |
| N2 slow delta power (F3) | 0.05 | 0.57 | N2 |
| N2 theta power (F3) | 0.04 | 0.59 | N2 |
| N2 fast delta power (F3) | 0.04 | 0.64 | N2 |
| N2 slow beta power (C3) | 0.03 | 0.68 | N2 |
| N2 slow beta power (C4) | 0.03 | 0.72 | N2 |
| N2 fast beta power (C4) | -0.03 | 0.70 | N2 |
| Spindles absolute power (C3) | 0.03 | 0.69 | N2 |
| N2 percentage | 0.03 | 0.72 | N2 |
| Spindles density (C3) | 0.02 | 0.83 | N2 |
| Spindles amplitude (C3) | 0.02 | 0.83 | N2 |
| Spindles absolute power (F3) | -0.02 | 0.81 | N2 |
| N2 duration | 0.02 | 0.80 | N2 |
| N2 slow delta power (C3) | -0.01 | 0.90 | N2 |
| N2 fast beta power (C3) | -0.01 | 0.91 | N2 |
| Spindles density (F3) | 0.01 | 0.91 | N2 |
| Spindles density (F4) | -0.01 | 0.89 | N2 |
| N2 fast delta power (C3) | 0.00 | 0.95 | N2 |
| N2 slow beta power (F3) | 0.00 | 1.00 | N2 |
| N2 slow delta power (C4) | 0.00 | 1.00 | N2 |
| N2 fast delta power (C4) | 0.00 | 0.99 | N2 |
| N2 fast beta power (F4) | 0.00 | 0.99 | N2 |
| Spindles density (C4) | 0.00 | 1.00 | N2 |

**Supplementary Table S3. Correlations of integrative resilience (latent score) with NREM stage 3 sleep microstructures.**

| **Feature** | **r coefficient** | **p value** | **Block** |
| --- | --- | --- | --- |
| N3 slow delta power (F4) | 0.28 | 0.00 | N3 |
| Slow waves negative peak amplitude (C4) | -0.25 | 0.00 | N3 |
| Slow waves negative peak amplitude (F4) | -0.23 | 0.00 | N3 |
| Slow waves density (F4) | 0.22 | 0.01 | N3 |
| N3 slow delta power (F3) | 0.19 | 0.01 | N3 |
| Slow waves negative peak amplitude (C3) | -0.20 | 0.02 | N3 |
| N3 theta power (O1) | -0.19 | 0.02 | N3 |
| N3 slow delta power (C3) | 0.17 | 0.03 | N3 |
| N3 fast delta power (O1) | -0.17 | 0.03 | N3 |
| N3 theta power (O2) | -0.18 | 0.03 | N3 |
| N3 slow beta power (O2) | -0.17 | 0.04 | N3 |
| Slow waves density (F3) | 0.17 | 0.04 | N3 |
| Slow waves negative peak amplitude (F3) | -0.18 | 0.03 | N3 |
| N3 slow delta power (C4) | 0.16 | 0.04 | N3 |
| N3 fast delta power (O2) | -0.16 | 0.05 | N3 |
| N3 fast beta power (O2) | -0.15 | 0.06 | N3 |
| N3 duration | 0.15 | 0.05 | N3 |
| N3 fast delta power (F4) | 0.15 | 0.07 | N3 |
| N3 alpha power (O1) | -0.14 | 0.09 | N3 |
| N3 slow beta power (O1) | -0.13 | 0.10 | N3 |
| N3 fast beta power (O1) | -0.13 | 0.11 | N3 |
| N3 alpha power (O2) | -0.13 | 0.11 | N3 |
| Slow waves duration (C3) | 0.14 | 0.09 | N3 |
| N3 percentage | 0.13 | 0.11 | N3 |
| N3 alpha power (F4) | 0.12 | 0.13 | N3 |
| Slow waves duration (F4) | 0.12 | 0.16 | N3 |
| Slow waves frequency (C3) | -0.11 | 0.17 | N3 |
| N3 slow sigma power (O1) | -0.10 | 0.20 | N3 |
| N3 theta power (F4) | 0.11 | 0.18 | N3 |
| N3 slow sigma power (O2) | -0.10 | 0.20 | N3 |
| N3 fast sigma power (O2) | -0.11 | 0.19 | N3 |
| Slow waves slope (F4) | 0.10 | 0.21 | N3 |
| Slow waves density (C4) | 0.10 | 0.25 | N3 |
| Slow waves duration (C4) | 0.09 | 0.25 | N3 |
| N3 slow sigma power (F4) | 0.09 | 0.29 | N3 |
| N3 fast sigma power (F4) | 0.09 | 0.27 | N3 |
| N3 fast sigma power (C4) | 0.09 | 0.28 | N3 |
| Slow waves positive peak amplitude (C4) | 0.09 | 0.30 | N3 |
| N3 fast delta power (F3) | 0.08 | 0.33 | N3 |
| N3 slow beta power (F4) | 0.08 | 0.32 | N3 |
| Slow waves frequency (F4) | -0.08 | 0.33 | N3 |
| N3 alpha power (F3) | 0.07 | 0.39 | N3 |
| N3 alpha power (C3) | 0.07 | 0.39 | N3 |
| N3 fast sigma power (O1) | -0.07 | 0.40 | N3 |
| N3 slow delta power (O2) | -0.07 | 0.39 | N3 |
| N3 alpha power (C4) | 0.07 | 0.39 | N3 |
| Slow waves frequency (C4) | -0.07 | 0.38 | N3 |
| Slow waves positive peak amplitude (F3) | 0.08 | 0.35 | N3 |
| N3 slow delta power (O1) | -0.07 | 0.41 | N3 |
| N3 fast sigma power (C3) | 0.07 | 0.42 | N3 |
| N3 fast beta power (F4) | 0.06 | 0.45 | N3 |
| Slow waves slope (F3) | 0.06 | 0.45 | N3 |
| Slow waves density (C3) | 0.06 | 0.47 | N3 |
| Slow waves duration (F3) | 0.06 | 0.50 | N3 |
| N3 theta power (C4) | 0.05 | 0.58 | N3 |
| Slow waves slope (C4) | 0.05 | 0.59 | N3 |
| Slow waves positive peak amplitude (C3) | 0.04 | 0.64 | N3 |
| N3 theta power (F3) | 0.03 | 0.71 | N3 |
| N3 theta power (C3) | 0.03 | 0.70 | N3 |
| N3 slow sigma power (F3) | 0.03 | 0.72 | N3 |
| N3 fast sigma power (F3) | 0.03 | 0.72 | N3 |
| N3 fast beta power (F3) | -0.03 | 0.72 | N3 |
| N3 slow sigma power (C4) | 0.03 | 0.69 | N3 |
| N3 slow sigma power (C3) | 0.02 | 0.77 | N3 |
| Slow waves frequency (F3) | -0.02 | 0.80 | N3 |
| N3 slow beta power (C4) | 0.02 | 0.85 | N3 |
| Slow waves slope (C3) | -0.01 | 0.87 | N3 |
| N3 fast delta power (C3) | -0.01 | 0.92 | N3 |
| N3 slow beta power (F3) | -0.01 | 0.93 | N3 |
| N3 slow beta power (C3) | 0.00 | 0.96 | N3 |
| N3 fast beta power (C3) | 0.00 | 0.96 | N3 |
| N3 fast beta power (C4) | 0.00 | 0.96 | N3 |
| N3 fast delta power (C4) | 0.00 | 0.99 | N3 |

**Supplementary Table S4. Correlations of integrative resilience (latent score) with REM sleep microstructures.**

| **Feature** | **r coefficient** | **p value** | **Block** |
| --- | --- | --- | --- |
| REM slow delta power (O1) | -0.17 | 0.03 | REM |
| REM fast delta power (O1) | -0.14 | 0.08 | REM |
| REM alpha power (O1) | -0.14 | 0.08 | REM |
| REM fast beta power (O1) | -0.15 | 0.07 | REM |
| REM slow delta power (O2) | -0.18 | 0.03 | REM |
| REM fast delta power (F4) | 0.17 | 0.03 | REM |
| REM fast delta power (O2) | -0.14 | 0.09 | REM |
| REM theta power (F4) | 0.17 | 0.04 | REM |
| REM theta power (O2) | -0.14 | 0.09 | REM |
| REM alpha power (O2) | -0.14 | 0.08 | REM |
| REM fast sigma power (F4) | 0.14 | 0.09 | REM |
| REM fast beta power (O2) | -0.16 | 0.05 | REM |
| REM duration | -0.14 | 0.08 | REM |
| REM percentage | -0.17 | 0.03 | REM |
| REM theta power (O1) | -0.13 | 0.11 | REM |
| REM slow beta power (O2) | -0.13 | 0.11 | REM |
| REM occipital aperiodic intercept | -0.13 | 0.12 | REM |
| REM slow delta power (C3) | -0.10 | 0.22 | REM |
| REM slow sigma power (O1) | -0.11 | 0.16 | REM |
| REM fast sigma power (O1) | -0.10 | 0.22 | REM |
| REM slow beta power (O1) | -0.10 | 0.20 | REM |
| REM fast beta power (F3) | -0.11 | 0.18 | REM |
| REM slow delta power (F4) | 0.12 | 0.15 | REM |
| REM alpha power (F4) | 0.10 | 0.21 | REM |
| REM slow sigma power (O2) | -0.12 | 0.15 | REM |
| REM fast sigma power (O2) | -0.12 | 0.14 | REM |
| REM slow beta power (F4) | 0.10 | 0.19 | REM |
| REM fast beta power (C4) | -0.10 | 0.20 | REM |
| REM frontal aperiodic slope | -0.11 | 0.17 | REM |
| REM frontal aperiodic intercept | 0.11 | 0.19 | REM |
| REM slow delta power (C4) | -0.10 | 0.24 | REM |
| REM fast sigma power (C3) | 0.08 | 0.30 | REM |
| REM theta power (C3) | 0.08 | 0.35 | REM |
| REM slow sigma power (C3) | 0.08 | 0.33 | REM |
| REM theta power (C4) | 0.08 | 0.35 | REM |
| REM central aperiodic slope | -0.08 | 0.33 | REM |
| REM slow sigma power (F4) | 0.07 | 0.37 | REM |
| REM fast sigma power (C4) | 0.07 | 0.37 | REM |
| REM fast delta power (C4) | 0.07 | 0.38 | REM |
| REM fast delta power (C3) | 0.06 | 0.42 | REM |
| REM slow sigma power (F3) | 0.06 | 0.46 | REM |
| REM fast sigma power (F3) | 0.06 | 0.46 | REM |
| REM fast beta power (C3) | -0.06 | 0.46 | REM |
| REM slow sigma power (C4) | 0.06 | 0.45 | REM |
| REM fast delta power (F3) | 0.05 | 0.53 | REM |
| REM theta power (F3) | 0.06 | 0.49 | REM |
| REM alpha power (C3) | 0.05 | 0.50 | REM |
| REM slow beta power (C3) | 0.05 | 0.53 | REM |
| REM fast beta power (F4) | -0.05 | 0.52 | REM |
| REM central aperiodic intercept | 0.05 | 0.52 | REM |
| REM alpha power (F3) | 0.05 | 0.57 | REM |
| REM alpha power (C4) | 0.04 | 0.59 | REM |
| REM slow beta power (C4) | 0.03 | 0.74 | REM |
| REM slow beta power (F3) | 0.02 | 0.77 | REM |
| REM slow delta power (F3) | -0.02 | 0.81 | REM |
| REM occipital aperiodic slope | -0.01 | 0.89 | REM |

**Supplementary Table S5. Comparisons of absolute power in frequency bands (log-transformed) in central region within NREM stage 2 sleep.**

| **Frequency** | **Electrode** | **Low integrative resilience** | **Moderate integrative resilience** | **High integrative resilience** |
| --- | --- | --- | --- | --- |
| Slow delta (0.1-1Hz) | (C3) | 1.67 (0.03) | 1.70 (0.02) | 1.64 (0.03) |
| Slow delta (0.1-1Hz) | (C4) | 1.68 (0.03) | 1.72 (0.02) | 1.66 (0.03) |
| Slow delta (0.1-1Hz) | (F3) | 1.58 (0.04) | 1.64 (0.03) | 1.60 (0.04) |
| Slow delta (0.1-1Hz) | (F4) | 1.53 (0.06) | 1.66 (0.03) | 1.65 (0.03) |
| Slow delta (0.1-1Hz) | (O1) | 1.75 (0.05) | 1.67 (0.04) | 1.49 (0.06) |
| Slow delta (0.1-1Hz) | (O2) | 1.77 (0.05) | 1.70 (0.04) | 1.50 (0.06) |
| Fast delta (1-3.5 Hz) | (C3) | 1.67 (0.02) | 1.69 (0.02) | 1.66 (0.03) |
| Fast delta (1-3.5 Hz) | (C4) | 1.66 (0.02) | 1.70 (0.02) | 1.66 (0.02) |
| Fast delta (1-3.5 Hz) | (F3) | 1.55 (0.04) | 1.58 (0.03) | 1.57 (0.03) |
| Fast delta (1-3.5 Hz) | (F4) | 1.51 (0.06) | 1.60 (0.03) | 1.62 (0.03) |
| Fast delta (1-3.5 Hz) | (O1) | 1.68 (0.06) | 1.58 (0.04) | 1.44 (0.06) |
| Fast delta (1-3.5 Hz) | (O2) | 1.70 (0.06) | 1.61 (0.04) | 1.45 (0.05) |
| Theta power (3.5-8Hz) | (C3) | 1.27 (0.03) | 1.30 (0.02) | 1.27 (0.03) |
| Theta power (3.5-8Hz) | (C4) | 1.26 (0.02) | 1.31 (0.02) | 1.29 (0.03) |
| Theta power (3.5-8Hz) | (F3) | 1.09 (0.04) | 1.12 (0.03) | 1.12 (0.04) |
| Theta power (3.5-8Hz) | (F4) | 1.06 (0.06) | 1.15 (0.03) | 1.19 (0.04) |
| Theta power (3.5-8Hz) | (O1) | 1.37 (0.05) | 1.28 (0.04) | 1.18 (0.05) |
| Theta power (3.5-8Hz) | (O2) | 1.40 (0.06) | 1.31 (0.04) | 1.19 (0.05) |
| Alpha power (8-12Hz) | (C3) | 0.82 (0.03) | 0.87 (0.03) | 0.83 (0.03) |
| Alpha power (8-12Hz) | (C4) | 0.82 (0.03) | 0.88 (0.02) | 0.85 (0.03) |
| Alpha power (8-12Hz) | (F3) | 0.67 (0.04) | 0.72 (0.03) | 0.71 (0.04) |
| Alpha power (8-12Hz) | (F4) | 0.66 (0.05) | 0.74 (0.03) | 0.77 (0.04) |
| Alpha power (8-12Hz) | (O1) | 0.90 (0.06) | 0.82 (0.04) | 0.72 (0.05) |
| Alpha power (8-12Hz) | (O2) | 0.92 (0.07) | 0.84 (0.04) | 0.72 (0.05) |
| Slow sigma (12-14Hz) | (C3) | 0.44 (0.03) | 0.45 (0.02) | 0.44 (0.03) |
| Slow sigma (12-14Hz) | (C4) | 0.43 (0.03) | 0.45 (0.02) | 0.46 (0.03) |
| Slow sigma (12-14Hz) | (F3) | 0.25 (0.03) | 0.26 (0.03) | 0.28 (0.04) |
| Slow sigma (12-14Hz) | (F4) | 0.27 (0.05) | 0.28 (0.03) | 0.34 (0.04) |
| Slow sigma (12-14Hz) | (O1) | 0.41 (0.05) | 0.32 (0.04) | 0.26 (0.05) |
| Slow sigma (12-14Hz) | (O2) | 0.41 (0.06) | 0.33 (0.04) | 0.24 (0.05) |
| Fast sigma (14-16Hz) | (C3) | 0.10 (0.03) | 0.09 (0.03) | 0.11 (0.04) |
| Fast sigma (14-16Hz) | (C4) | 0.09 (0.03) | 0.10 (0.02) | 0.13 (0.04) |
| Fast sigma (14-16Hz) | (F3) | -0.12 (0.03) | -0.14 (0.02) | -0.09 (0.04) |
| Fast sigma (14-16Hz) | (F4) | -0.12 (0.04) | -0.12 (0.03) | -0.03 (0.04) |
| Fast sigma (14-16Hz) | (O1) | 0.08 (0.05) | 0 (0.04) | -0.04 (0.05) |
| Fast sigma (14-16Hz) | (O2) | 0.10 (0.07) | 0.01 (0.03) | -0.06 (0.05) |
| Slow beta (16-24Hz) | (C3) | 0.21 (0.03) | 0.19 (0.03) | 0.20 (0.03) |
| Slow beta (16-24Hz) | (C4) | 0.20 (0.03) | 0.19 (0.02) | 0.22 (0.03) |
| Slow beta (16-24Hz) | (F3) | 0.09 (0.03) | 0.05 (0.02) | 0.10 (0.04) |
| Slow beta (16-24Hz) | (F4) | 0.08 (0.04) | 0.08 (0.02) | 0.15 (0.03) |
| Slow beta (16-24Hz) | (O1) | 0.26 (0.05) | 0.17 (0.04) | 0.12 (0.05) |
| Slow beta (16-24Hz) | (O2) | 0.31 (0.07) | 0.19 (0.04) | 0.10 (0.05) |
| Fast beta (14-32Hz) | (C3) | -0.17 (0.03) | -0.20 (0.03) | -0.20 (0.03) |
| Fast beta (14-32Hz) | (C4) | -0.17 (0.03) | -0.20 (0.02) | -0.19 (0.03) |
| Fast beta (14-32Hz) | (F3) | -0.25 (0.04) | -0.31 (0.02) | -0.27 (0.04) |
| Fast beta (14-32Hz) | (F4) | -0.25 (0.04) | -0.29 (0.02) | -0.23 (0.04) |
| Fast beta (14-32Hz) | (O1) | -0.13 (0.04) | -0.23 (0.03) | -0.28 (0.05) |
| Fast beta (14-32Hz) | (O2) | -0.07 (0.07) | -0.21 (0.03) | -0.29 (0.04) |

Note: Descriptive data were shown as Mean (SE).

**Supplementary Table S6. Comparisons of absolute power in frequency bands (log-transformed) in NREM stage 3 sleep.**

| **Frequency** | **Electrode** | **Low integrative resilience** | **Moderate integrative resilience** | **High integrative resilience** |
| --- | --- | --- | --- | --- |
| Slow delta (0.1-1Hz) | (C3) | 2.43 (0.03) | 2.51 (0.02) | 2.51 (0.04) |
| Slow delta (0.1-1Hz) | (C4) | 2.46 (0.03) | 2.54 (0.02) | 2.54 (0.04) |
| Slow delta (0.1-1Hz) | (F3) | 2.32 (0.05) | 2.44 (0.03) | 2.49 (0.04) |
| Slow delta (0.1-1Hz) | (F4) | 2.30 (0.05) | 2.47 (0.03) | 2.53 (0.04) |
| Slow delta (0.1-1Hz) | (O1) | 2.52 (0.07) | 2.49 (0.05) | 2.4 (0.07) |
| Slow delta (0.1-1Hz) | (O2) | 2.52 (0.06) | 2.51 (0.05) | 2.39 (0.06) |
| Fast delta (1-3.5 Hz) | (C3) | 2.34 (0.03) | 2.3 (0.02) | 2.3 (0.03) |
| Fast delta (1-3.5 Hz) | (C4) | 2.33 (0.03) | 2.33 (0.02) | 2.3 (0.03) |
| Fast delta (1-3.5 Hz) | (F3) | 2.24 (0.04) | 2.24 (0.03) | 2.28 (0.05) |
| Fast delta (1-3.5 Hz) | (F4) | 2.23 (0.04) | 2.26 (0.03) | 2.32 (0.04) |
| Fast delta (1-3.5 Hz) | (O1) | 2.31 (0.08) | 2.16 (0.05) | 2.01 (0.06) |
| Fast delta (1-3.5 Hz) | (O2) | 2.29 (0.08) | 2.17 (0.05) | 2.01 (0.06) |
| Theta power (3.5-8Hz) | (C3) | 1.56 (0.03) | 1.54 (0.02) | 1.54 (0.03) |
| Theta power (3.5-8Hz) | (C4) | 1.55 (0.02) | 1.56 (0.02) | 1.55 (0.03) |
| Theta power (3.5-8Hz) | (F3) | 1.43 (0.03) | 1.42 (0.02) | 1.44 (0.04) |
| Theta power (3.5-8Hz) | (F4) | 1.42 (0.04) | 1.44 (0.03) | 1.49 (0.04) |
| Theta power (3.5-8Hz) | (O1) | 1.65 (0.06) | 1.52 (0.04) | 1.42 (0.05) |
| Theta power (3.5-8Hz) | (O2) | 1.66 (0.06) | 1.55 (0.04) | 1.44 (0.05) |
| Alpha power (8-12Hz) | (C3) | 0.95 (0.04) | 0.96 (0.02) | 0.96 (0.04) |
| Alpha power (8-12Hz) | (C4) | 0.93 (0.04) | 0.98 (0.02) | 0.96 (0.04) |
| Alpha power (8-12Hz) | (F3) | 0.87 (0.04) | 0.88 (0.03) | 0.91 (0.05) |
| Alpha power (8-12Hz) | (F4) | 0.85 (0.05) | 0.90 (0.03) | 0.96 (0.05) |
| Alpha power (8-12Hz) | (O1) | 0.97 (0.07) | 0.85 (0.04) | 0.75 (0.05) |
| Alpha power (8-12Hz) | (O2) | 0.98 (0.08) | 0.87 (0.04) | 0.76 (0.05) |
| Slow sigma (12-14Hz) | (C3) | 0.45 (0.04) | 0.43 (0.02) | 0.45 (0.04) |
| Slow sigma (12-14Hz) | (C4) | 0.43 (0.04) | 0.44 (0.02) | 0.46 (0.04) |
| Slow sigma (12-14Hz) | (F3) | 0.29 (0.04) | 0.27 (0.03) | 0.31 (0.05) |
| Slow sigma (12-14Hz) | (F4) | 0.28 (0.05) | 0.30 (0.03) | 0.37 (0.05) |
| Slow sigma (12-14Hz) | (O1) | 0.35 (0.06) | 0.26 (0.04) | 0.22 (0.06) |
| Slow sigma (12-14Hz) | (O2) | 0.35 (0.07) | 0.28 (0.04) | 0.2 (0.05) |
| Fast sigma (14-16Hz) | (C3) | 0.01 (0.04) | -0.02 (0.02) | 0.00 (0.04) |
| Fast sigma (14-16Hz) | (C4) | 0.00 (0.04) | 0.00 (0.02) | 0.02 (0.04) |
| Fast sigma (14-16Hz) | (F3) | -0.20 (0.04) | -0.24 (0.02) | -0.20 (0.04) |
| Fast sigma (14-16Hz) | (F4) | -0.21 (0.05) | -0.22 (0.02) | -0.15 (0.04) |
| Fast sigma (14-16Hz) | (O1) | -0.03 (0.05) | -0.11 (0.03) | -0.16 (0.05) |
| Fast sigma (14-16Hz) | (O2) | 0.00 (0.07) | -0.10 (0.03) | -0.18 (0.05) |
| Slow beta (16-24Hz) | (C3) | 0.05 (0.03) | 0.03 (0.02) | 0.02 (0.03) |
| Slow beta (16-24Hz) | (C4) | 0.05 (0.03) | 0.05 (0.02) | 0.04 (0.03) |
| Slow beta (16-24Hz) | (F3) | -0.07 (0.04) | -0.09 (0.02) | -0.06 (0.03) |
| Slow beta (16-24Hz) | (F4) | -0.07 (0.04) | -0.06 (0.02) | -0.03 (0.03) |
| Slow beta (16-24Hz) | (O1) | 0.11 (0.05) | 0.03 (0.03) | -0.05 (0.04) |
| Slow beta (16-24Hz) | (O2) | 0.17 (0.08) | 0.05 (0.03) | -0.05 (0.04) |
| Fast beta (14-32Hz) | (C3) | -0.33 (0.03) | -0.35 (0.02) | -0.35 (0.03) |
| Fast beta (14-32Hz) | (C4) | -0.33 (0.03) | -0.33 (0.02) | -0.34 (0.03) |
| Fast beta (14-32Hz) | (F3) | -0.42 (0.04) | -0.44 (0.02) | -0.42 (0.04) |
| Fast beta (14-32Hz) | (F4) | -0.43 (0.04) | -0.41 (0.02) | -0.38 (0.04) |
| Fast beta (14-32Hz) | (O1) | -0.27 (0.05) | -0.34 (0.03) | -0.4 (0.04) |
| Fast beta (14-32Hz) | (O2) | -0.2 (0.08) | -0.31 (0.03) | -0.4 (0.04) |

**Note:** Descriptive data were shown as Mean (SE).

**Supplementary Table S7. Comparisons of absolute power in frequency bands (log-transformed) in REM sleep.**

| **Frequency** | **Electrode** | **Low integrative resilience** | **Moderate integrative resilience** | **High integrative resilience** |
| --- | --- | --- | --- | --- |
| Slow delta (0.1-1Hz) | (C3) | 1.39 (0.03) | 1.35 (0.02) | 1.31 (0.03) |
| Slow delta (0.1-1Hz) | (C4) | 1.39 (0.03) | 1.35 (0.02) | 1.32 (0.03) |
| Slow delta (0.1-1Hz) | (F3) | 1.35 (0.04) | 1.32 (0.02) | 1.34 (0.03) |
| Slow delta (0.1-1Hz) | (F4) | 1.29 (0.06) | 1.33 (0.03) | 1.36 (0.03) |
| Slow delta (0.1-1Hz) | (O1) | 1.41 (0.06) | 1.3 (0.05) | 1.17 (0.06) |
| Slow delta (0.1-1Hz) | (O2) | 1.45 (0.06) | 1.33 (0.05) | 1.18 (0.06) |
| Fast delta (1-3.5 Hz) | (C3) | 1.29 (0.03) | 1.29 (0.02) | 1.29 (0.03) |
| Fast delta (1-3.5 Hz) | (C4) | 1.28 (0.03) | 1.29 (0.02) | 1.30 (0.03) |
| Fast delta (1-3.5 Hz) | (F3) | 1.18 (0.04) | 1.16 (0.03) | 1.21 (0.04) |
| Fast delta (1-3.5 Hz) | (F4) | 1.13 (0.06) | 1.19 (0.03) | 1.26 (0.04) |
| Fast delta (1-3.5 Hz) | (O1) | 1.31 (0.05) | 1.21 (0.04) | 1.12 (0.05) |
| Fast delta (1-3.5 Hz) | (O2) | 1.34 (0.06) | 1.25 (0.04) | 1.14 (0.05) |
| Theta power (3.5-8Hz) | (C3) | 1.01 (0.03) | 1.03 (0.02) | 1.03 (0.03) |
| Theta power (3.5-8Hz) | (C4) | 1.01 (0.03) | 1.03 (0.02) | 1.04 (0.03) |
| Theta power (3.5-8Hz) | (F3) | 0.88 (0.05) | 0.86 (0.03) | 0.91 (0.04) |
| Theta power (3.5-8Hz) | (F4) | 0.83 (0.06) | 0.9 (0.03) | 0.97 (0.04) |
| Theta power (3.5-8Hz) | (O1) | 1.07 (0.05) | 0.98 (0.04) | 0.9 (0.05) |
| Theta power (3.5-8Hz) | (O2) | 1.11 (0.06) | 1.01 (0.04) | 0.90 (0.05) |
| Alpha power (8-12Hz) | (C3) | 0.53 (0.03) | 0.55 (0.03) | 0.53 (0.04) |
| Alpha power (8-12Hz) | (C4) | 0.54 (0.02) | 0.56 (0.02) | 0.55 (0.04) |
| Alpha power (8-12Hz) | (F3) | 0.36 (0.05) | 0.35 (0.03) | 0.39 (0.05) |
| Alpha power (8-12Hz) | (F4) | 0.36 (0.05) | 0.38 (0.03) | 0.44 (0.04) |
| Alpha power (8-12Hz) | (O1) | 0.76 (0.05) | 0.66 (0.04) | 0.56 (0.05) |
| Alpha power (8-12Hz) | (O2) | 0.80 (0.06) | 0.70 (0.05) | 0.57 (0.05) |
| Slow sigma (12-14Hz) | (C3) | -0.11 (0.03) | -0.09 (0.03) | -0.08 (0.04) |
| Slow sigma (12-14Hz) | (C4) | -0.10 (0.03) | -0.07 (0.02) | -0.07 (0.03) |
| Slow sigma (12-14Hz) | (F3) | -0.26 (0.04) | -0.27 (0.03) | -0.22 (0.04) |
| Slow sigma (12-14Hz) | (F4) | -0.24 (0.06) | -0.23 (0.03) | -0.17 (0.04) |
| Slow sigma (12-14Hz) | (O1) | 0.11 (0.05) | 0.02 (0.04) | -0.05 (0.05) |
| Slow sigma (12-14Hz) | (O2) | 0.14 (0.07) | 0.06 (0.04) | -0.05 (0.05) |
| Fast sigma (14-16Hz) | (C3) | -0.23 (0.03) | -0.21 (0.03) | -0.20 (0.04) |
| Fast sigma (14-16Hz) | (C4) | -0.22 (0.03) | -0.20 (0.02) | -0.18 (0.03) |
| Fast sigma (14-16Hz) | (F3) | -0.36 (0.04) | -0.37 (0.02) | -0.32 (0.04) |
| Fast sigma (14-16Hz) | (F4) | -0.38 (0.04) | -0.33 (0.03) | -0.26 (0.04) |
| Fast sigma (14-16Hz) | (O1) | -0.08 (0.05) | -0.15 (0.04) | -0.21 (0.05) |
| Fast sigma (14-16Hz) | (O2) | -0.04 (0.07) | -0.12 (0.04) | -0.22 (0.05) |
| Slow beta (16-24Hz) | (C3) | 0.24 (0.03) | 0.23 (0.03) | 0.27 (0.04) |
| Slow beta (16-24Hz) | (C4) | 0.24 (0.03) | 0.24 (0.02) | 0.27 (0.04) |
| Slow beta (16-24Hz) | (F3) | 0.16 (0.04) | 0.13 (0.03) | 0.2 (0.04) |
| Slow beta (16-24Hz) | (F4) | 0.14 (0.04) | 0.17 (0.03) | 0.25 (0.04) |
| Slow beta (16-24Hz) | (O1) | 0.29 (0.05) | 0.2 (0.04) | 0.16 (0.06) |
| Slow beta (16-24Hz) | (O2) | 0.34 (0.07) | 0.24 (0.04) | 0.14 (0.06) |
| Fast beta (14-32Hz) | (C3) | 0.00(0.04) | -0.06 (0.03) | -0.05 (0.05) |
| Fast beta (14-32Hz) | (C4) | 0.00 (0.04) | -0.06 (0.03) | -0.05 (0.04) |
| Fast beta (14-32Hz) | (F3) | -0.02 (0.05) | -0.11 (0.03) | -0.08 (0.04) |
| Fast beta (14-32Hz) | (F4) | -0.03 (0.06) | -0.07 (0.03) | -0.03 (0.05) |
| Fast beta (14-32Hz) | (O1) | -0.04 (0.05) | -0.16 (0.04) | -0.22 (0.06) |
| Fast beta (14-32Hz) | (O2) | 0.00 (0.08) | -0.13 (0.05) | -0.24 (0.06) |

**Note:** Descriptive data were shown as Mean (SE).

**Supplementary Table S8. Comparisons of slow waves characteristics across resilience groups.**

| **Variables** | **Electrode** | **Low integrative resilience** | **Moderate integrative resilience** | **High integrative resilience** |
| --- | --- | --- | --- | --- |
| Slow waves density | (C3) | 6.80 (0.71) | 7.06 (0.54) | 6.98 (0.65) |
| Slow waves density | (C4) | 6.75 (0.66) | 7.62 (0.56) | 7.38 (0.60) |
| Slow waves density | (F3) | 5.13 (0.71) | 6.21 (0.53) | 7.52 (0.79) |
| Slow waves density | (F4) | 5.15 (0.77) | 6.76 (0.57) | 7.97 (0.77) |
| Slow wave slope | (C3) | 398.24 (7.94) | 390.25 (6.54) | 388.98 (9.55) |
| Slow wave slope | (C4) | 392.46 (7.87) | 396.84 (6.36) | 391.82 (8.02) |
| Slow wave slope | (F3) | 386.78 (9.54) | 381.90 (7.07) | 394.69 (12.55) |
| Slow wave slope | (F4) | 381.9 (10.64) | 385.23 (7.75) | 397.89 (11.45) |
| Slow waves negative peak amplitude | (C3) | -61.63 (1.12) | -63.17 (0.64) | -64.18 (0.87) |
| Slow waves negative peak amplitude | (C4) | -61.28 (1.22) | -63.94 (0.65) | -65.11 (0.91) |
| Slow waves negative peak amplitude | (F3) | -59.7 (1.28) | -62.16 (0.89) | -63.03 (0.93) |
| Slow waves negative peak amplitude | (F4) | -58.74 (1.41) | -62.48 (0.98) | -63.72 (1.16) |
| Slow wave positive peak amplitude | (C3) | 50.70 (1.06) | 50.42 (0.73) | 50.47 (0.89) |
| Slow wave positive peak amplitude | (C4) | 50.73 (0.90) | 51.68 (0.74) | 51.49 (0.82) |
| Slow wave positive peak amplitude | (F3) | 47.73 (1.01) | 48.31 (0.69) | 49.01 (0.95) |
| Slow wave positive peak amplitude | (F4) | 46.92 (1.15) | 48.61 (0.75) | 49.78 (0.92) |

Note: Descriptive data were shown as Mean (SE).

**Supplementary Table S9. Comparisons of melatonin levels in sequences across resilience groups.**

| **Variable** | **Low integrative resilience** | **Moderate integrative resilience** | **High integrative resilience** |
| --- | --- | --- | --- |
| DLMO time | 22:56 (0:14) | 22:43 (0:09) | 22:19 (0:14) |
| Melatonin level (pmol/L) |  |  |  |
| T1 | 4.65 (0.00) | 4.65 (0.00) | 5.08 (0.19) |
| T2 | 4.69 (0.03) | 7.27 (1.98) | 4.79 (0.08) |
| T3 | 4.72 (0.05) | 5.79 (0.69) | 4.83 (0.08) |
| T4 | 4.66 (0.01) | 5.24 (0.25) | 5.02 (0.13) |
| T5 | 4.66 (0.00) | 7.49 (1.40) | 5.77 (0.46) |
| T6 | 4.65 (0.00) | 7.10 (0.99) | 6.60 (1.00) |
| T7 | 5.65 (0.49) | 10.20 (1.67) | 8.98 (1.56) |
| T8 | 10.53 (2.21) | 15.76 (2.65) | 14.70 (2.32) |
| T9 | 19.04 (3.93) | 24.51 (3.30) | 29.14 (4.20) |
| T10 | 31.51 (6.07) | 39.28 (4.25) | 46.09 (5.92) |
| T11 | 44.76 (7.58) | 56.38 (5.09) | 63.36 (7.65) |
| T12 | 57.46 (8.60) | 74.69 (6.54) | 81.90 (8.67) |
| T13 | 69.22 (9.23) | 89.13 (8.00) | 95.34 (8.17) |
| T14 | 84.47 (9.39) | 111.75 (11.12) | 109.47 (9.05) |
| T15 | 88.56 (9.29) | 118.68 (11.12) | 120.14 (10.74) |
| T16 | 102.39 (9.56) | 126.70 (11.91) | 130.40 (10.62) |
| T17 | 108.50 (10.1) | 137.51 (12.81) | 139.82 (12.05) |

**Note:** Descriptive data were shown as Median (IQR).

**Supplementary Table S10. Correlations of integrative resilience (latent score) with circadian rhythm (rest activity pattern and DLMO time).**

| Feature | Spearman’s rho coefficient | p value | Block |
| --- | --- | --- | --- |
| DLMO time | -0.20 | 0.02 | Circadian |
| Acrophase | -0.14 | 0.09 | Circadian |
| Onset time of the active period | -0.14 | 0.08 | Circadian |
| Interdaily stability | 0.04 | 0.60 | Circadian |
| Intradaily variability | -0.07 | 0.39 | Circadian |
| Relative amplitude | -0.03 | 0.70 | Circadian |
| Activity of the rest period | 0.02 | 0.86 | Circadian |
| Activity of the active period | -0.01 | 0.87 | Circadian |
| Onset time of the rest period | -0.07 | 0.40 | Circadian |

**Supplementary text**

**Protocol of at-home and in-laboratory assessments of Dim Light Melatonin Onset**

**The at-home assessment:** Our research staff visited participants’ home to set up the ambulatory PSG on the night prior to the assessment. During this visit, participants were provided with detailed instructions and demonstrations of dim lighting. The indoor dim light environment was prepared by closing all blinds and curtains, dimming all indoor lights, and minimizing the light intensity of electronic media (e.g., cell phones, televisions, computers, and iPads) to the lowest level possible for participants’ reference. When giving the dim light environment demonstration, light intensity was controlled below 20 lux, measured at eye level, in the direction of gaze, approximately 30-40 cm from the eye using the TENMARS TM-201 Light Meter. Participants were instructed to roll up sleeves or wear short sleeve shirts to ensure that the light sensor of the actigraphy would not be covered. Participants were instructed to remain awake and in the same position as much as possible under dim light, and collected salivary samples every 30 minutes. Each participant was provided with a home salivary collection kit, which included 17 collection tubes with prepared label informing the correSpindles onding sequence (e.g., “T1” for 6 hr before HBT, “T2” for 5.5 hr before HBT), a pack of straws, a pair of blue-light blocker glasses, several removable ice packs and an insulation box. Night lights were provided to participants to assist in dimming the room, and blue light blockers glasses were also provided to participants only for exceptional circumstances that participants had to enter a common area where lighting was unavoidable (e.g., washroom). Details of the modified at-home methodology for measuring was published in our recent study[1].

To ensure sampling time compliance, participants were instructed to send text messages to research staff about the actual sample collecting time immediately after they had collected the salivary samples. Our research staff would also send reminder messages to participants if they did not self-report sample collections within 5 minutes. After collecting all salivary samples, participants were instructed to immediately store them in the freezer (-20°C - -4°C) until researchers collected them back the next day.

**The in-laboratory assessment:** The in-laboratory was performed in the Laboratory of Sleep Sciences at the University of Hong Kong. The laboratory is equipped with a controllable light system for each bedroom. Participants were invited to arrive at least 15 min before the assessment started. When the laboratory assessment started, participants were instructed to remain awake and in the same position as much as possible under dim light (< 20 lux, at eye level, in the direction of gaze, 30-40 cm from eyes). During the assessment period, participants were prompted by laboratory staff to give salivary samples every 30 minutes. The salivary sample collection protocol was the same as the at-home protocol. After collection, all samples were immediately stored in -20°C freezer.
